# Vaccination as personal public-good provision

**DOI:** 10.1101/2022.04.21.22274110

**Authors:** J. Lucas Reddinger, Gary Charness, David Levine

**Affiliations:** Department of Economics, Purdue University, West Lafayette, Indiana, 47907; Department of Economics, University of California, Santa Barbara, 93106; Department of Economics, University of Wisconsin-La Crosse, 54601; Haas School of Business, University of California, Berkeley, 94720

**Keywords:** social preferences, prosocial behavior, vaccination, behavioral public health, health policy, COVID-19, SARS-CoV-2, B1.617.2, public good, altruism, efficiency, equity, experimental economics

## Abstract

Vaccination against infectious diseases has both private and public benefits. We study whether social preferences—concerns for the well-being of other people—are associated with one’s decision regarding vaccination. We measure these social preferences for 549 online subjects with a public-good game and an altruism game. To the extent that one gets vaccinated out of concern for the health of others, contribution in the public-good game is analogous to an individual’s decision to obtain vaccination, while our altruism game provides a different measure of altruism, equity, and efficiency concerns. We proxy vaccine demand with how quickly a representative individual voluntarily took the initial vaccination for COVID-19 (after the vaccine was widely available). We collect COVID-19 vaccination history separately from the games to avoid experimenter-demand effects. We find a strong result: Contribution in the public-good game is associated with greater demand to voluntarily receive a first dose, and thus also to vaccinate earlier. Compared to a subject who contributes nothing, one who contributes the maximum ($4) is 58% more likely to obtain a first dose voluntarily in the four-month period that we study (April through August 2021). In short, people who are more pro-social are more likely to take a voluntary COVID-19 vaccination. Behavior in our altruism game does not predict vaccination. We recommend further research on the use of pro-social preferences to help motivate individuals to vaccinate for other transmissible diseases, such as the flu and HPV.

## 1. Introduction

Vaccines prevent an estimated 6 million deaths annually (Rodrigues and Plotkin 2020). Beyond morbidity and mortality prevention, workers are more productive, economic growth greater, and equity enhanced. Despite these private and public benefits, vaccine hesitancy continues to pose a challenge. Addressing COVID-19 vaccination hesitancy remains a top priority in public health (Krause, Gruber, and Offit 2021). While largely vaccine-preventable, seasonal influenza annually causes 200,000 to 800,000 hospitalizations and 20,000 to 60,000 deaths in the U.S. (CDC 2020a). Further, clusters of measles cases have recurred in countries that once were free of the disease, while greater HPV vaccination is a cost-effective means to improve health (Maldonado, O’Leary, and Hotez 2022; Chesson et al. 2011).

While the strongest personal motivation for vaccination is most likely the direct benefits to one’s own health, the decision to vaccinate could be influenced by a variety of pro-social concerns, such as helping one’s community. One may wish to vaccinate to help attain herd immunity (reflecting altruism, a desire for social efficiency, or some combination). In addition, one may wish to do one’s fair share (a preference for equity) if many other people in their community are vaccinated. The fact that people do not simply maximize their own material payoffs may have important consequences for public health.

Immunity to communicable disease is a public good (Althouse, Bergstrom, and Bergstrom 2010). When one vaccinates against many communicable diseases (e.g., the flu, measles, HPV, COVID-19), one incurs personal health benefits, and vaccination lowers transmission rates to others (Wissing et al. 2019; CDC 2021b, 2020b, 2021a). As with other public goods, a free-riding problem exists in vaccination. One has a high incentive to vaccinate if no one else is vaccinated against a prevalent communicable disease, but a much lower incentive if most other people are vaccinated. While immunized individuals are still susceptible to infection, broader vaccination reduces community transmission.

For example, Loeb et al. (2010) find that vaccinating school children and adolescents for influenza protects non-immunized community members (attributed to decreased transmission in schools). Similarly, White (2021) finds that each flu vaccination among the general population of the United States generates $63 of benefits from reduced mortality and $87 of benefits from work hours gained. Vaccination against HPV provides significant private benefits to both women and men, and the vaccination of each group helps protect the other (Elbasha and Dasbach 2010). Regarding COVID-19 vaccination, Oliu-Barton et al. (2022) estimate that French vaccination certificate regulations saved an average of €685 of economic output per vaccination.

People also can have social preferences, where they deliberately sacrifice money or other resources to help (or hurt) others, to establish equity or efficiency, or to increase the economic surplus for the group. As one indicator of the importance of social preferences, Americans donated over $500 billion to charitable organizations in 2020. Although subjects in laboratory experiments are frequently concerned with the payoffs of others, some question whether this behavior is generally present in the field. We implemented two incentivized games to measure social preferences.

Our public-good game endowed each online participant in anonymous groups of four with $4. Each person chose how much of their $4 to contribute to a public good, while retaining the remainder. We then doubled the group’s total contribution and distributed it evenly to all participants. Thus, each player receives back half of what they contributed, while providing benefits for the group. The socially-optimal choice would be for each person to contribute $4 to the pot, so that each person earns $8; however, free-riding is the dominant individual strategy to maximize material payoffs.

To the extent that one gets vaccinated out of concern for the well-being of others, voluntary vaccination is analogous to this public-good game. One may (perhaps reasonably) believe that vaccination is not directly worthwhile, considering only one’s private benefits and costs; however, one certainly benefits from the vaccination of others.

Participants also played an altruism game, in which the respondent chose how much money (between $0 and $6) another randomly-selected participant would receive, while the respondent receives $4 regardless of the choice. This altruism game provides a measure of monetarily-costless altruism. Note that a sole preference for altruism or efficiency suggests a choice of $6, while a sole preference for fairness suggests a choice of $4.

In a separate survey, we asked participants to report being vaccinated for COVID-19 and, if so, the date of their first dose. COVID-19 vaccination is an ideal case study—widespread and expedient uptake was sought, which allows us to use one’s delay to vaccinate over a four-month period as a proxy for vaccine demand. To avoid any contamination and potential experimenter-demand effects, we conducted the COVID-19 vaccine survey about one month before the social-preference games, using a different researcher identity.

We sampled individuals who reported being unvaccinated during an earlier survey in April 2021. Participants then completed a COVID-19 survey in mid-August and incentivized games in September and October. To control for local factors such as availability of vaccines and social norms, we use national data to estimate the daily vaccination (hazard) rate of a first dose among the unvaccinated for each U.S. county from April 10 to August 13, 2021. We model the vaccination hazard rate, including the daily number of doses available per capita in a state, daily county-by-date COVID-19 case rates, test positivity rates, vaccination rates, and a time trend. These public data account for substantial heterogeneity by obtaining county-by-date predicted first-dose vaccination rate.

Finally, we model the behavior of our participants. We assume that an unvaccinated individual’s demand for a first dose (their hazard rate of vaccination) is proportional to their pro-social preferences (as measured in the games) and the hazard rate for a representative unvaccinated person in their county on each date. We separately model vaccination that participants attribute to a mandate at work or school, because pro-social preferences should have a considerably lesser role in this case.^5^

We consider the correlation between social preferences and vaccination behavior; prior research has studied intent to receive a hypothetical vaccination (often for a flu virus), as well as COVID-19 prophylactic behavior (e.g., masking, handwashing). Studies on this topic have shown some conflicting results, as described in our literature review. While there is some evidence (e.g., Milkman et al. 2022) that nudges or directives help to induce more pro-social behavior, some studies have failed to demonstrate this (e.g., Campos-Mercade et al. 2021a). We measure social preferences from behavior in incentivized games designed to elicit social preferences, a relatively uncommon methodology in the vaccination literature. We completely separate social-preference elicitation from measurement of vaccination behavior, as we conduct these sessions a month apart under different researcher identities.^6^

We are one of the first to study the relationship between contributions in a public-good game and actual vaccination behavior (as opposed to self-reported intent to vaccinate).^7^ We capture the vaccination behavior of our participants during the first four crucial months that the COVID-19 vaccines were available to the general public of the United States.^8^ Finally, participants who report having taken the vaccine due to a mandate at work or school likely have different motivation; our model permits these motivations to differ. In total, we use an innovative methodology to offer unique findings on the association between trait-level social preferences and actual COVID-19 vaccination in the field.

We find clear results: Compared to a subject who contributes nothing in the public-good game, one who contributes the maximum ($4) is on average 58% more likely to obtain a first dose voluntarily in the four-month period that we study (April through August 2021). That is, contribution in the public-good game is positively associated with voluntary vaccination. However behavior in our altruism game is not predictive of vaccination. This particular combination of results suggests that fairness or equity concerns are the channels through which pro-social preferences predict voluntary vaccination.

While our evidence is consistent with the hypothesis that contribution in a public-good game is correlated with voluntary vaccination against COVID-19, these results do not necessarily translate directly to policy advice on COVID-19 vaccination efforts. To the extent that our results are relevant to vaccination for transmissible diseases writ large, pro-social motivation may be harnessed to increase vaccination for the flu (as by Milkman et al. 2022), HPV, or measles.

## 2. Prior literature

The literature on pro-social motivations for vaccination comprises a body of mixed evidence. Some of these studies find an association between pro-social preferences and vaccination, while others do not. Most studies use non-incentivized survey questions to measure social preferences, while only a few use incentivized games. Most studies ask participants to report only their intent to vaccinate against a real infectious disease, usually the flu (see Böhm and Betsch (2022) for a review). Our unique contribution is our combination of incentivized games to measure social preferences, actual vaccination timing in the field, and a comprehensive set of robustness checks (for example, our incentivized games are highly predictive, even when controlling for predicted vaccination hazard or attitudes about COVID-19 vaccination).

Many studies have sought to link social considerations to vaccination decisions, but in total, this body of evidence is mixed. For example, Hershey et al. (1994) survey patients at a student health clinic about intent to vaccinate against a hypothetical influenza disease. While altruism, free-riding, and herding all motivate the intent to vaccinate, there is no evidence that stressing societal benefits of vaccination increases this intent. Betsch et al. (2017) test whether explaining the concept of herd immunity to participants increases their reported intent to vaccinate in fictitious scenarios. Contrary to the research hypothesis, this intervention increases reported intent to vaccinate among residents of Western countries, but not those of more-collectivist Eastern countries. Amin et al. (2017) find no association between intent to vaccinate and self-reported attitudes toward fairness and pro-sociality. Betsch and Böhm (2018) study the interaction of an intervention that explains the social benefits of vaccination with one’s self-reported attitudes toward fairness and pro-sociality, finding no interactive effect. Böhm, Betsch, and Korn (2016) find marginal evidence that subjects more cooperative in dictator games were more likely to choose to “vaccinate” in a framed laboratory game. In the field, Milkman et al. (2022) find that text messages sent to encourage flu vaccination increase uptake. Message content that emphasizes the protection of others is beneficial; however, some messages without social concerns perform better, and other messages worse. Given the general lack of consensus regarding pro-social vaccination motives, our contribution is potentially valuable.

In the specific context of pro-social motives for COVID-19 vaccination, most evidence fails to find a association. One exception is Pfattheicher, Petersen, and Böhm (2021), who find that knowledge about the social benefits of COVID-19 vaccination is associated with greater intent to vaccinate among respondents residing in the United Kingdom (U.K.). These authors also find that an intervention that explains the social benefits of vaccination increases self-reported intent to vaccinate. The remaining evidence is largely negative. Freeman et al. (2021) conduct a study of COVID-19 vaccination intentions among 15,014 U.K. residents solicited from a variety of sources, including TV, radio, and mail campaigns. These authors find no effect of information provision regarding the societal benefits of COVID-19 vaccination. In fact, the authors find that providing strongly-hesitant respondents with information on the *private* benefit of vaccination increases stated intent to vaccinate for COVID-19, but that the addition of information about collective benefits reduces intent to vaccinate. Banker and Park (2020) find that prosocial messages (“protect your community”) were in fact significantly less effective than those using a self-focused frame (“protect yourself”) in eliciting click-throughs to official CDC recommendations in a large field experiment conducted on Facebook during the critical initial weeks of the COVID-19 outbreak in the United States. Finally, Campos-Mercade et al. (2021a) do not find that a pro-social nudge (that stresses that the COVID-19 vaccine protects others) increases vaccination uptake (both studies use administrative vaccination records). These mixed results echo those from studies conducted pre-COVID-19; a consensus view has clearly not yet emerged.^9^ We contribute to this discussion using lab-in-the-field incentivized games along with vaccination timing as a proxy for vaccination hazard.

Much of the evidence for social preferences in economics has come from simple games in laboratory experiments. There are concerns that laboratory measures of social preferences may not translate into consistent behaviors outside of the lab (Gneezy and List 2006; List 2006; Levitt and List 2007). At the same time, field behavior often matches experimental behavior (e.g., Falk and Heckman 2009; Bellemare, Bissonnette, and Kröger 2014).^10^ In combining incentivized games with actual vaccination timing in the field, we further validate laboratory measures of social preferences.

Thus, we ask whether the choices made in two games with modest-but-real stakes inform us to any degree about actual vaccination behavior in the field. We find that it does.

## 3. Methodology

### 3.1. Survey methods

We begin with an online “Survey about COVID-19 vaccination” (Session I) that solicits demographic characteristics and asks if and when the respondant received a first-dose of a COVID-19 vaccine, for $1 compensation. We focus solely on an individual’s first dose for comparability in both costs and timing, given that some vaccines required two doses to achieve a complete vaccination, while other vaccines available at the time only required a single dose.^11^ If the respondent reports having received a first dose, we ask in which month, then whether it was early (days 1-10), in the middle (days 11-20), or late (days 21-31) in that month.

We conduct a separate survey (Session II) weeks later, using a different researcher identity to collect incentivized measures of pro-social preferences (thus avoiding contamination from the survey about vaccination). This second survey pays $1 in addition to payment from one randomly-selected game. Each game is referred to as a scenario, administered using the strategy method, and played in the same order by all participants.

Each participant first played our altruism game (“Game A”), in which the respondent receives $4 and chooses how much another respondent will receive, from $0 to $6, without affecting the respondent’s own earnings. A coin toss chooses one of the players’ endowment choices to be implemented for the other.

Participants then played our public-good game (“Game B”), in which each person chooses how much of their $4 to contribute to a public good, while retaining the remainder. We randomly match groups of four, double the group’s total contribution, and distribute this amount evenly among the group members.

We thought that Game A would capture monetarily-costless altruistic preferences, as altruism in the public-good game comes at personal monetary cost. But we considered the public-good game as the most analogous to vaccination against a transmissible disease, since the benefits to others of vaccination seems to reflect public-good provision more than simple altruism. Accordingly, for the public-good game, we prepared a sequence of ten detailed instructional graphics; Game A only had a single page of textual instructions, in part due to this game’s relative simplicity.

For each participant we randomly select a game for payment; we randomly pair subjects with the same game selection; we then pay each participant the outcome associated with their and their counterpart’s choices.^12^

### 3.2. Statistical methods

An unvaccinated individual faces a continual decision of whether or not to take a first vaccine dose. However, upon receiving a first dose, the individual no longer faces this decision (and thus no behavior may be observed). Accordingly, survival analysis is the appropriate framework for this decision process (and standard in this literature).^13^ The hazard rate is the probability of receiving a first dose conditional on being unvaccinated. Given a relatively small sample of individuals, we control for many influences that affect the demand for and supply of a first dose.

We begin by modeling this hazard rate using public daily county data to account for heterogeneity across time and geography in factors affecting the supply of and demand of vaccines. A linear lasso procedure selects among candidate covariates (described in the following section and listed in Table 1) and predicts vaccination hazard in U.S. county *c* in state *s* on date *t*. This estimate is the daily probability of receiving a first dose, conditional on never having received a dose. We use this estimated hazard rate as a control variable in our main regression.

**Table 1:** Variables used in the linear lasso to predict dose hazard rate*_c_*_,*s*,*t*_.

| Variable |
| --- |
| Time trend |
| Indicator of B1.617.2 (“Delta”) having emerged in the U.S. |
| Indicator of state governor being Republican |
| Vaccine doses available per capita<sup>a</sup> |
| Cases per capita<sup>b,c</sup> |
| Percentage of tests positive<sup>b,c</sup> |
| Maximum cases per capita to date in the county<sup>c</sup> |
| Maximum test positivity rate to date in the county<sup>c</sup> |
| Percent of residents having received a first dose<sup>a,c</sup> |
| Percent of residents having completed series<sup>a,c,d</sup> |

<sup>a</sup> Seven-day moving average.
<sup>b</sup> Seven-day change.
<sup>c</sup> These variables were additionally included with a one-week lag; they are also included as interacted with the indicator of whether Delta had yet emerged in the U.S.
<sup>d</sup> Having received one dose of the Johnson & Johnson’s Janssen JNJ-78436735 vaccine or two doses of either the Pfizer-BioNTech BNT162b2 or Moderna mRNA-1273 vaccine.

Turning to our participant data, our model considers whether vaccination is voluntary or if the respondents said it was mandated by their employer or school. We use a proportional hazard model with two covariates—predicted hazard rate for each participant’s county *c* in state *s*on date *t* and each participant’s contribution in our public-good game. We then test our primary hypothesis that pro-social preferences, as measured in the public-good game, predict the decision to receive a first dose of a COVID-19 vaccine.

### 3.3. Modeling the hazard of a first dose for each county-day

Our main control variation is the hazard rate of receiving a first dose, estimated for each county and date.

An individual faces costs in getting vaccinated; these include time spent finding an available vaccine dose and traveling to the clinic. If vaccine doses are extremely scarce, an individual may spend a considerable amount of time searching for an available appointment and may be willing to travel farther. At the margin, these costs likely translate to a lower first-dose hazard rate. To capture the effect of vaccine scarcity, we consider the number of vaccine doses that are available per person on each date. The CDC (2022c) report the daily number of doses delivered to every state, as well as the number administered, both per capita. We consider the difference to be the number of doses available per capita, of which we take a seven-day moving average.

We also consider county-level factors that influence an individual’s private benefit from vaccination. This benefit increases with a higher local case rate, so we include the number of new cases per capita recorded in a county over the last seven days. Because limited testing can underestimate case rates, we also include the local percentage of tests reported positive over the last seven days. One’s decision may also be affected by salient data such as the worst cases and test-positivity rates experienced locally. For example, if there have been severe local rates, the consequences of high community transmission may be more salient. So, we also include the maximal test-positivity and cases-per-capita rates to date for each county. All county-level case and test-positivity data come from the CDC (2022d).

One also incurs greater benefits from vaccination when fewer individuals in one’s community are vaccinated. To capture this, we include the proportion of county residents who have received a first dose and the proportion who have completed their vaccination series, both as seven-day moving averages. These county-level vaccination data come from the CDC (2022b). Due to differences with B1.617.2 (“Delta”), we include an indicator of whether the variant had yet emerged in the U.S., as reported by the CDC (2022a). We also interact this indicator variable with the previous variables, to capture differences in the costs and benefits for this variant, and further include the variables above with a one-week lag. Public policy regarding testing, test reporting, vaccination, incentives, and mandates are all highly-politicized in the United States, so we include an indicator of whether the state governor is a member of the Republican party (Kaiser Family Foundation 2021). We also include a time trend.

Finally, the regressand of interest is the hazard rate for a first dose in the county on that date. We construct the probability that an adult becomes vaccinated, conditional on being an unvaccinated adult within county *c* in state *s* at time *t*, as

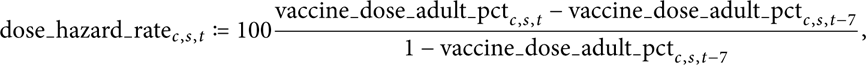

where vaccine dose adult pct*_c_* _,*s*, *t*_ is a seven-day moving average of the percentage of adults receiving a first dose in county *c* on day *t*, again from the CDC (2022b).

We employ a linear lasso to select among the variables described above, which are also listed in Table 1. We use the resulting model to predict 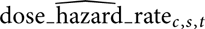 for every county on each date. We merge these predictions to our participants by county-date. We use approximated geolocation of each participant’s zip code, some of which span multiple counties. If multiple counties match, we use the county with the largest share of residential addresses in the given zip code, according to data from Housing and Urban Development (2021). The inclusion of these predicted hazard rates absorbs a substantial amount of variance from our regressions of vaccination behavior reported by our participants.

### 3.4. Modeling the hazard rate of vaccination in our sample

We now analyze the timing of first doses in our participant sample. We consider two possible causes of vaccination for any individual: voluntary or compelled by a mandate. Suppose an individual decides on April 1st to wait to take her first dose voluntarily. In June, her employer gives her two weeks to take a first dose to retain employment; she complies. In this sense, she was at risk for both non-mandated and mandated vaccination, until she took her first dose under pressure. Had she not been vaccinated under a mandate, she may have vaccinated voluntarily regardless in August, but we cannot observe this counterfactual behavior. Accordingly, her behavior regarding nonmandated vaccination is censored once she becomes vaccinated due to a mandate. More formally, each individual’s behavior is characterized by two choices—the time at which she would take a first dose without a mandate, and the time at which she would take a first dose under a mandate. We observe only the first of these events (or neither). When we observe one event, behavior regarding the other event is censored in future time periods.

We base our statistical approach on our hypothesis. Because we are primarily interested in pro-social motivation to vaccinate, we focus on voluntary vaccination. With our first approach— analysis of the cause-specific hazard—we consider individual behavior in a hypothetical world without any vaccination mandates. This approach is appropriate to investigate whether pro-social preferences are associated with an individual’s decision to vaccinate in the absence of a mandate. To this end, we use a Cox proportional-hazards model and censor individuals who become vaccinated due to a mandate at work or school (Pintilie 2007).^14^ We specify first-dose hazard not attributed to a mandate as

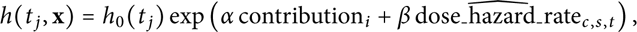

where *h*_0_ (*t _j_*) is the baseline hazard (the hazard given covariate values of zero), endowment*_i_* ∈ {0, 1, …, 6} is the choice of individual *i* in the altruism game, contribution*_i_* ∈ {0, 0.5, …, 4} is the choice of individual *i* in the public-good game, and 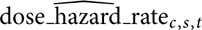 is the predicted first-dose hazard in that county on that date (as described in the previous section).^15^ We asked respondents to report the date of their first dose in bins of roughly 10 days with midpoints {*t _j_*} to facilitate responses to our survey.

We do not apply the Cox regression to vaccination attributed to a mandate. This method would only be appropriate to investigate vaccination attributed to a mandate in a hypothetical world without voluntary vaccination. Our primary approach of studying the cause-specific (non-mandated) first-dose hazard assumes that vaccination due to a mandate is uninformative. However, individuals who attribute their vaccination to a mandate may possess characteristics that would also inform their decision of when to vaccinate without mandate. In Appendix C, we present similar results with a competing-hazards model (Fine and Gray 1999).

## 4. Results

### 4.1. Description of the sample

We initially recruited U.S. residents from Prolific’s participant pool April 7 – 27, 2021, using a single-question survey asking their vaccination status.^16^ Prolific is a marketing firm that maintains a pool of survey respondents. We over-sampled participants who had already self-identified with Prolific as Black, Latinx, Hispanic, a parent, or politically conservative; we sought these characteristics for a separate study (Reddinger, Levine, and Charness 2022).

We conducted Session I on August 12 and 13, 2021. We asked about participants’ demographic characteristics, COVID-19 vaccination history, and attitudes toward COVID-19 vaccination, receiving 601 responses. Of these respondents, 549 participated in Session II, which was available from September 11 to October 13, 2021, and contained the incentivized games and questions about pro-social attitudes. We randomly chose one game for payment for each participant; we then randomly assigned groups for each game. The participants’ choices determined payments in the games, which we made within 24 hours of participation.^17^

Of our 549 participants, three did not indicate whether they had been vaccinated. Of the remainder, 306 reported being unvaccinated and 240 reported being vaccinated. Of the vaccinated respondents, seven were unsure of the date or did not report one. Our analysis thus generally reports on the 539 respondents who participated in the two games and who also provided their vaccination status and, if so, the approximate date of their first COVID-19 dose.

Figure 1 describes our sample, reporting behavior in each game, as well as participant age, education, and vaccination timings. Our respondents are younger than the general population of the United States. Most of our participants have at least some college education; many have a college degree. Roughly one-third of our sample identifies as Republican, one-third Democrat, and one-third either “Independent” or “other.” Of vaccinated subjects, 15.4% (37 of 240) attributed their vaccination to a mandate at work, at school, for travel, or to attend social events or restaurants.^18^ Recall that in our altruism game, a participant could endow their counterpart with $0 to $6, receiving $4 regardless. The mean and median choice was $4, mode $6, and a substantial mass at $0.

**Figure 1:**
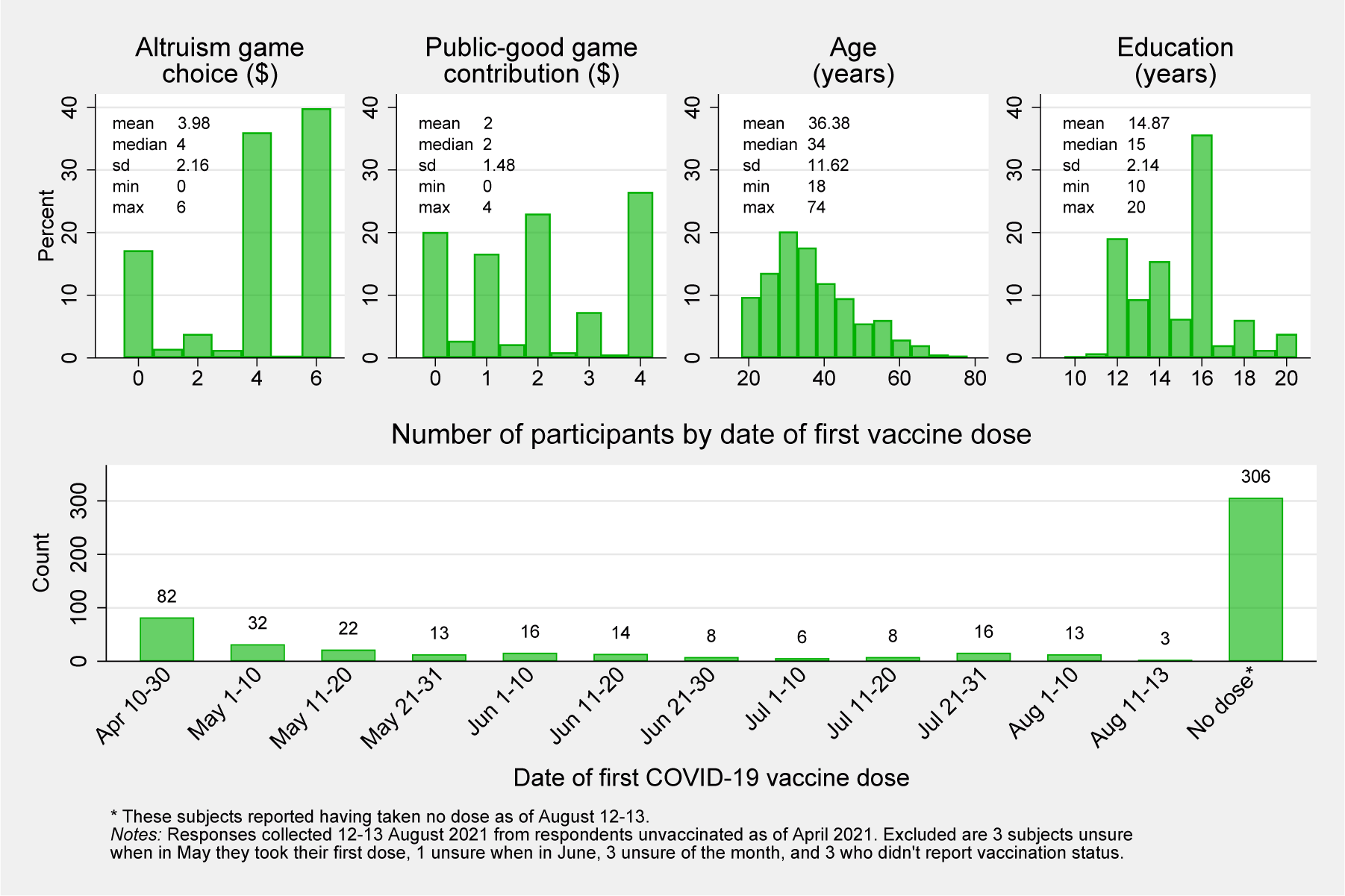
Participant characteristics.

In our public-good game, we gave participants a $4 bonus from which they could contribute any amount to a pot that doubled and was then split equally among their group of four randomly-matched participants. The mean and median contribution was $2; we note that many experiments find initial public-good contributions average 40–60% of the social optimum (Ledyard 1995). Table 2 presents correlation coefficients between behavior in the two social-preference games, survey responses about social considerations of COVID-19 vaccination, and survey responses about social attitudes. Choices in the altruism game are generally not significantly associated with social attitude responses, suggesting it may not prove a useful measure of social preferences. Contribution in the public-good game is significantly associated with nearly all social attitude responses, suggesting its potential usefulness. We thus conclude that behavior in the public-good game is valid as a measure of social preferences. Choices in neither game are associated with either two other-regarding COVID-19 survey questions (Table 2, Panel B), suggesting that the public-good game may capture nuanced preferences (such as for efficiency or fairness) that go beyond the vaccination’s ability to protect one’s own friends and family. We further explore this in Appendix B.

**Table 2:** Correlation coefficients between game behavior and other-regarding survey responses.

|  | Mean | S.D. | Game A |  | Game B |  |
| --- | --- | --- | --- | --- | --- | --- |
| | | | N | $\rho$ | N | $\rho$ |
| PANEL A: BEHAVIOR IN GAMES DESIGNED TO MEASURE SOCIAL PREFERENCES (SESSION II, PART I) |  |  |  |  |  |  |
| Altruism game choice (\$, Game A) | 3.98 | 2.16 | 547 | | | |
| Contribution in public-good game (\$, Game B) | 2.00 | 1.48 | 546 | 0.201*** | | |
| PANEL B: SURVEY OF SOCIAL CONCERNS OF COVID-19 VACCINATION<sup>1</sup> (SESSION I) |  |  |  |  |  |  |
| Getting vaccinated would help protect the health of my friends and family. | 3.48 | 1.50 | 547 | 0.013 | 547 | 0.048 |
| My friends or family want me to get vaccinated. | 3.53 | 1.32 | 546 | −0.003 | 546 | −0.004 |
| PANEL C: SURVEY RESPONSES REGARDING SOCIAL ATTITUDES<sup>1</sup> (SESSION II, PART II) |  |  |  |  |  |  |
| Helping others is usually a waste of time. | 1.64 | 0.83 | 546 | −0.073<sup>+</sup> | 546 | −0.203*** |
| When given the opportunity, I enjoy aiding others who are in need. | 4.23 | 0.80 | 547 | 0.060 | 547 | 0.132** |
| Helping friends and family is one of the great joys in life. | 4.27 | 0.87 | 546 | 0.060 | 546 | 0.141*** |
| It feels wonderful to assist others in need. | 4.32 | 0.79 | 547 | 0.039 | 547 | 0.150*** |
| Volunteering to help someone is very rewarding. | 4.25 | 0.87 | 545 | 0.047 | 545 | 0.117** |
| I dislike giving directions to strangers who are lost. | 2.14 | 1.15 | 546 | −0.021 | 546 | −0.056 |
| Unless they are part of my family, helping the elderly isn't my responsibility. | 2.04 | 1.06 | 545 | 0.058 | 545 | −0.121** |
| If the person in front of me in the check-out line at a store was a few cents short, I would pay the difference. | 4.23 | 1.04 | 547 | 0.090* | 547 | 0.193*** |

|  | Mean | S.D. | Game A |  | Game B |  |
| --- | --- | --- | --- | --- | --- | --- |
| | | | N | $\rho$ | N | $\rho$ |
| I plan to donate my organs when I die with the hope that they will help someone else live. | 3.42 | 1.48 | 547 | 0.081<sup>+</sup> | 547 | 0.187*** |
| Helping people does more harm than good because they come to rely on others and not themselves. | 1.95 | 1.01 | 546 | -0.137** | 546 | -0.182*** |

<sup>+</sup> $p < 0.10$ , \* $p < 0.05$ , \*\* $p < 0.01$ , \*\*\* $p < 0.001$ <sup>1</sup> Likert scale of 5 “Strongly agree,” 4 “Somewhat agree,” 3 “Neither agree nor disagree,” 2 “Somewhat disagree,” and 1 “Strongly disagree.”

Table 3 shows the average choice in each game by the individual’s vaccination status as of August 2021. Figure 2 plots vaccination over time for different levels of contribution in the public-good game, showing that vaccination differs by choices in each of the games. These Kaplan-Meier curves make no modeling assumptions; they are purely descriptive representations of our data. For the altruism game we partition choices into three subsets, $0 to 3.5 (less than one’s own payoff), $4 (equal to), and $4.5 to 6 (greater than). We partition behavior in the public-good game into three sets of three adjacent choices each. Figure 2 makes it clear that vaccination timing is empirically meaningful, further motivating our use of survival analysis; this information is lost in Table 3, which only compares vaccination status at one arbitrary date.

**Table 3:** Behavior in each game by vaccination status.

|  | Vaccinated,<br>no mandate | Vaccinated,<br>mandated | Not<br>vaccinated |
| --- | --- | --- | --- |
| Altruism game choice (\$, Game A) | 4.03<br>(2.15) | 3.89<br>(2.11) | 3.97<br>(2.17) |
| Contribution in public-good game (\$, Game B) | 2.15<br>(1.48) | 1.62<br>(1.24) | 1.95<br>(1.50) |
| Observations | 203 | 37 | 304 |

**Figure 2:**
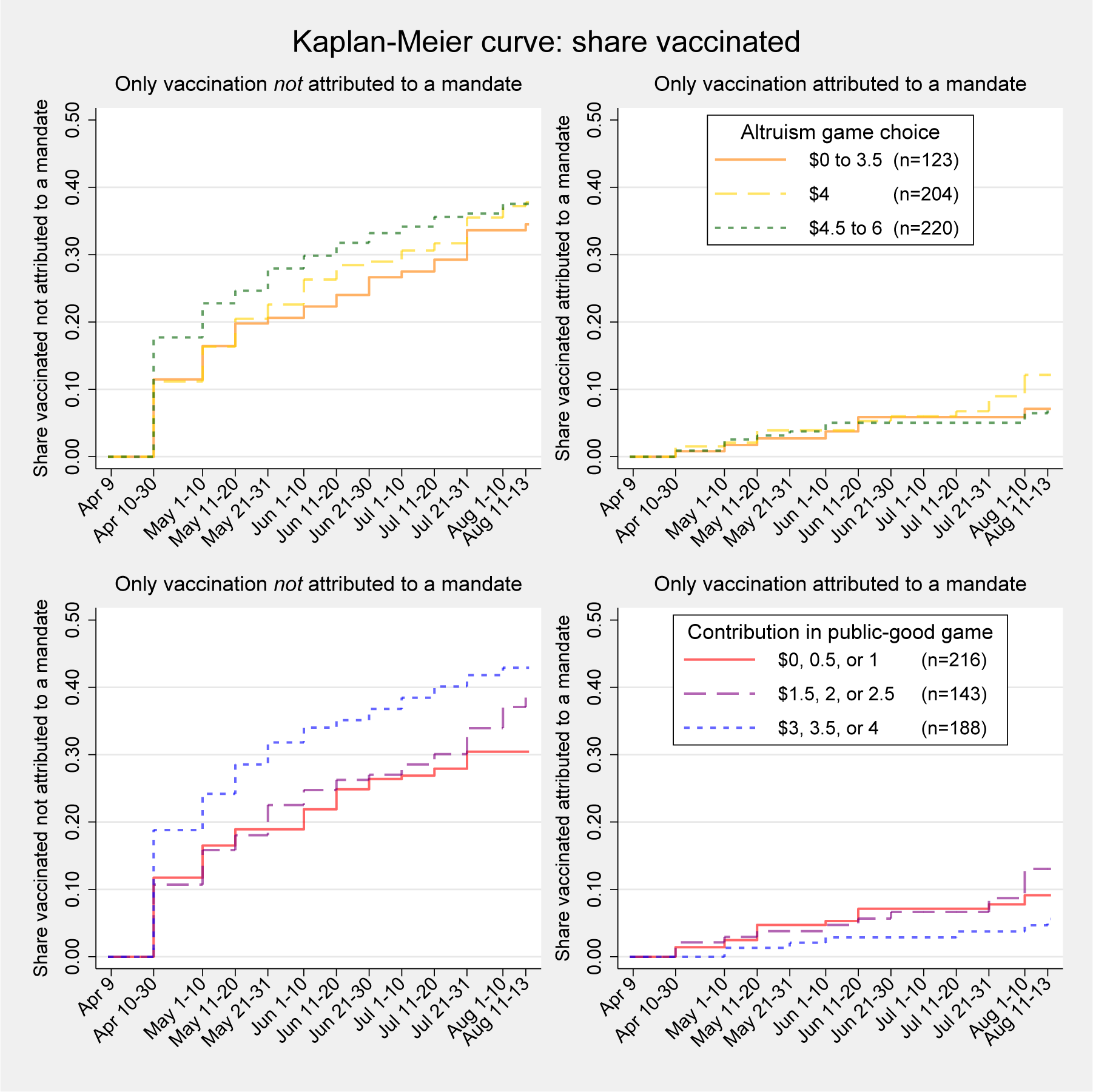
Vaccination over time by choice in each game.

### 4.2. Inference

We now consider our primary hypothesis that choices in the two incentivized games—our measures of pro-social preferences—are positively associated with vaccination. We consider results from the Cox model, with regression results shown in Table 4. Recall that this approach focuses on vaccination not attributed to a mandate in a hypothetical world without any vaccination mandates. We do not find an effect regarding the altruism game (Columns 1 and 2). Meanwhile, an additional dollar contribution in the public-good game is associated with a 12.1% greater daily hazard of vaccination not attributed to a mandate. This result stands without use of any control variables (Table 4, Column 3). An individual contributing the full $4 in the game is 1.58 times as likely to vaccinate (not due to a mandate) at any time (Column 3, *p* = 0.024; Column 4, *p* = 0.020) than one contributing nothing, as also depicted in Figure 3.^19^ This provides strong support for our hypothesis that pro-social preferences, as measured in the public-good game, predict voluntary vaccination.

**Table 4:** Cox regressions of vaccination hazard (not attributed to a mandate)

|  | (1) | (2) | (3) | (4) |
| --- | --- | --- | --- | --- |
| Endowment to other participant (\$, Game A) | 1.010<br>(0.035) | 1.009<br>(0.035) | | |
| Contribution in public-good game (\$, Game B) | 1.116*<br>(0.057) | 1.123*<br>(0.059) | 1.121*<br>(0.056) | 1.127*<br>(0.058) |
| Predicted county-by-date first-dose hazard rate |  | 1.048<br>(0.053) |  | 1.049<br>(0.053) |
| Subjects | 538 | 538 | 539 | 539 |
| Observations (subject-by-date) | 5468 | 5468 | 5482 | 5482 |

Notes: Coefficients exponentiated (hazard ratios). Robust standard errors in parentheses. Efron method used for ties. Vaccination attributed to a mandate (at work or school) is censored.
<sup>+</sup> $p < 0.10$ , \* $p < 0.05$ , \*\* $p < 0.01$ , \*\*\* $p < 0.001$ .

**Figure 3:**
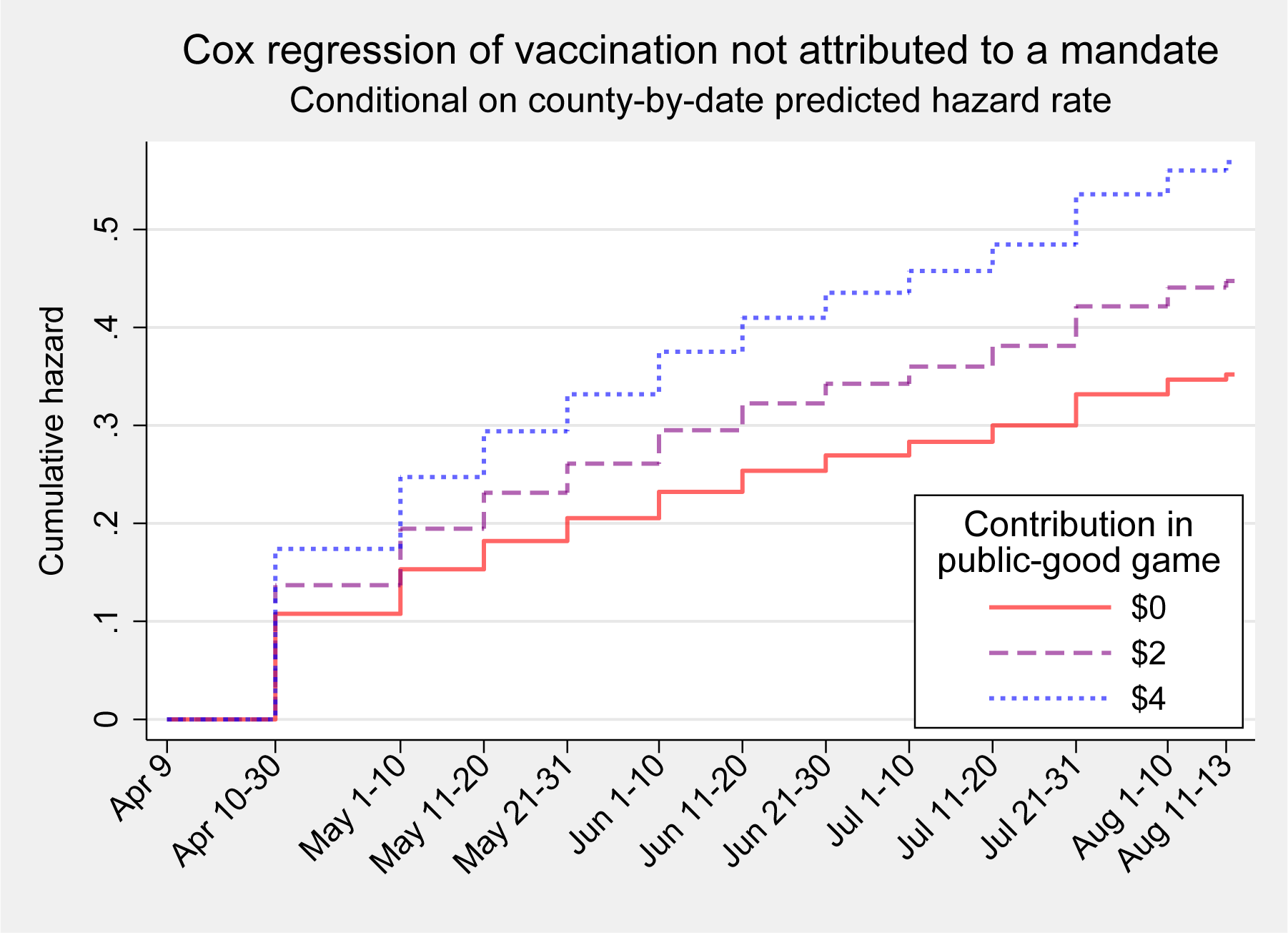
Cox regression of vaccination hazard not attributed to a mandate.

The coefficients for game choices give the additional vaccination (hazard rate or ratio) per dollar, conditional on the covariates. When we use no other covariates (as in Table 4, Columns 1 and 3), the baseline vaccination hazard is estimated entirely from our own sample. When we use the predicted value for each county-by-date (Columns 2 and 4), the baseline hazard is calculated for our sample, but conditional on what would be predicted for the average person in that county on that date. This demonstrates that our results are robust to the best predictions for our subjects by using relevant public-health data.^20^

Appendix A presents the remainder of our pre-registered analysis plan, which considers comprehension checks and the inclusion of additional individual co-variates, to which our results are robust. Appendix B presents significant results using a Tobit model, an OLS model, and different standard errors specifications; it also validates the proportional-hazards assumption of the Cox regressions. Appendix C presents results using a competing-hazards model.

We felt that our public-good game was a good match for the decision to vaccinate, given the efficiency factor in the public-good game (contributions were doubled) and the indirect social benefits (see Charness and Rabin 2002; Althouse, Bergstrom, and Bergstrom 2010). However, we included our altruism game to test whether monetarily-costless altruism might be driving any results. In fact, choices in the altruism game did not predict vaccination, while the effect of public-good contribution remains statistically significant. This suggests that altruism itself is not driving the results.

In the public-good game, an individual with a prevailing preference for equity would act as they believe others will act in the game. Otherwise, preferences for altruism or efficiency suggest a maximal ($4) contribution. In the altruism game, the own-payoff is $4, so equity would dictate a choice of $4, while altruism and efficiency suggest $6. Taken together, individuals with a strong preference for equity may be more likely to vaccinate, choose $4 in the public-good game, and choose $4 in the altruism game (causing it to be less predictive). We thus conclude that equity is an important motivator in the decision to vaccinate.

In total, the public-good game, like vaccination, uniquely permits an outcome that is both socially efficient and equitable, an outcome lacking in the altruism game. A combination of fairness and a desire for social efficiency plausibly drives both contribution in our public-good game and early vaccination against COVID-19.

## 5. Conclusion

We ask whether social preferences—concerns for the well-being of other people—influence one’s decision regarding vaccination. We measure social preferences using two incentivized games, a public-good game and an altruism game. Our sample consisted of unvaccinated U.S. residents as of April 2021, and we collected vaccination history through August 2021. People who are more pro-social in the public-good game are more likely to get a voluntary vaccination (and to get it earlier). Compared to a subject who contributes nothing, one who contributes the maximum ($4) is 58% more likely to obtain a first dose voluntarily in the four-month period that we study (April through August 2021). The altruism game is not predictive, suggesting that equity or efficiency concerns prevail as motives to vaccinate.

Whether policy-makers can harness pro-social preferences to increase vaccination remains an open question. Prior experimental evidence on hypothetical vaccination finds mixed success (see Section 2). However, vaccination in the field differs from hypothetical vaccination, and COVID-19 vaccination is particularly hampered by politics and misinformation. We separately consider how our findings may inform efforts to increase vaccination for influenza, HPV, and COVID-19.

Influenza vaccination, like COVID-19 vaccination, confers less protection to the elderly compared to younger people, while the disease poses far greater health risks to the elderly (CDC 2021a; DiazGranados et al. 2014). Epidemiologists have suggested that vaccinating school children would yield greater social benefits than vaccinating the elderly, because the decrease in transmission among children in schools attributed to the immunization of another child is more beneficial than immunizing another elderly person (Medlock and Galvani 2009; Galvani, Reluga, and Chapman 2007). Many public schools in the U.S. offer flu vaccine clinics, for which parental consent must first be obtained. As one example of a simple intervention, when obtaining informed consent, clinic administrators could provide parents with information sheets that emphasize how vaccination of their own children saves the lives of elders in their community—perhaps a child’s own grandparents.

HPV vaccination is currently recommended by the CDC to both men and women of ages 11 to 26. For HPV infections acquired in 2018, the lifetime cost for all women of ages 15–59 in the U.S. was $543 million, while for men the costs totaled $205 million (Chesson et al. 2021). Men clearly have less private incentive to vaccinate for HPV than do women, yet one’s vaccination benefits the public at large. By 2018, 55% of females of ages 18 – 21 had taken a first HPV vaccine dose, while only 34% of men had done so (Chen et al. 2021). When recommending HPV vaccination, a primary care provider could stress to a young male patient that his own vaccination could prevent his future wife acquiring cervical cancer, the fourth-leading cause of cancer among women (Hirth 2019).

Campaigns promoting vaccination should carefully test specific language and messaging. Our results hint that equity and efficiency concerns may prevail in decisions to vaccinate. An appeal to equity may prove effective, such as “Together we can all vaccinate against HPV and dramatically reduce cancer risk among women and men.” Alternately, a message that stresses efficiency might be particularly persuasive, such as “When children get a quick flu shot at school, we save the lives of elders in our community.”

Similar interventions suggest less potential upside for COVID-19 vaccination promotion. Many pro-social individuals have already vaccinated for COVID-19 (as seen in our study); further, many of the holdouts are likely more galvanized against vaccination for COVID-19 than for the flu. Field interventions involving pro-social motivators might be worthwhile to test; however, by July 2022 (when we are writing), those unvaccinated for COVID-19 may be more concerned with personal risks than public benefits (e.g., Freeman et al. 2021).

Monetary incentives have the possibility to increase vaccination, but policy-makers ought to consider potential adverse effects. Low monetary incentives can crowd-out pro-social behavior (Gneezy and Rustichini 2000). Low financial incentives may reduce the “warm glow” felt by individuals who vaccinate out of concern for others. Further, low monetary incentives may also lead individuals to believe that social benefits are lower than they really are, also potentially decreasing pro-social vaccination. Evidence typically supports the effectiveness of monetary incentives to vaccinate for COVID-19, but not always.^21^

Our study finds marginally-significant evidence that vaccination attributed to a mandate is associated with weaker pro-social preferences (see Appendix C). We would expect individuals with weaker pro-social preferences to respond more strongly to private incentives, including workplace and school mandates. However, even if this weak result holds, it does not imply that mandates will necessarily be effective. Additional widespread mandates may reduce the warm glow felt by individuals who vaccinate out of concern for others. However, Karaivanov et al. (2021) find evidence that COVID-19 vaccination mandates are very effective overall, limiting such a concern. Schmelz and Bowles (2021) note that the implementation of mandates may increase COVID-19 vaccination hesitancy. Indeed, mandatory policies can foster mistrust and backlash (Falk and Kosfeld 2006).

Given the challenges with existing approaches to increase COVID-19 vaccination uptake, policy-makers and researchers may yet experiment with means to harness pro-social motives, but ought also address individuals’ beliefs regarding private benefits and risks. To the extent that pro-social motivation has been exhausted for COVID-19 vaccination, carrots such as incentives and sticks such as mandates have proven fruitful.

Pro-social motives have considerable potential as a useful tool to increase vaccination for many infectious diseases, such as seasonal influenza, HPV, and measles.

## Data Availability

All data produced are available online at https://doi.org/10.17605/OSF.IO/K6WE2.

## Acknowledgements

We thank four anonymous referees and the editors for their comments and Owen O’Donnell for his helpfulness.

## Authorship contributions

Reddinger: Conceptualization, methodology, formal analysis, writing, project administration, software, validation, investigation, resources, data curation, visualization. Charness: Conceptualization, methodology, formal analysis, writing, project administration. Levine: Conceptualization, methodology, formal analysis, writing, project administration, funding acquisition.

## Appendix A. Remaining pre-registered analysis

As first summarized in Section 3.1, our pre-analysis plan (Reddinger, Charness, and Levine 2021) specified three points of analysis:

(a) Test whether behavior in $4-endowed incentivized games predicts self-reported vaccination;
(b) test comprehension using two games; and
(c) include regression analyses with covariates from Session I.

Section 4.2 reported our primary results which uses the games to predict vaccination. In this section we include results for the remaining two points (b) and (c) of our pre-analysis plan.

### A.1. Comprehension checks

Concerned that our online participants may quickly click through the survey, we pre-specified the use of two games as comprehension checks. We thought that these games might be needed to cut through noisy data. Section 4.2 presented strong results without subsampling on our comprehension checks. We prefer those results, with minimal sampling decisions to lessen any concern of multiple hypothesis testing. Nevertheless, for completeness, this section considers such subsampling.

Participants played two games styled after the “Sigma-dominated” games found in Gupta, Rigotti, and Wilson (2021), which these authors use as a diagnostic for inattention. Our GRW-sigma-dominated-style games, with payoffs scaled to match the $4 outcomes in our social-preference games, are shown in Table 5. In these games, the self-interested option is aligned with the socially-efficient option. We expect participants to play Green if they understand the games. However, note that these games were presented to participants in a terse manner, whereas the two social-preference games were explained in an intuitive manner using graphics when helpful. Thus the usefulness of these games as comprehension checks may be limited.

**Table 5:** Payoffs for comprehension check Games C and D.

| Game C |  | Other |  | Game D |  | Other |  |
| --- | --- | --- | --- | --- | --- | --- | --- |
|  |  | Green | Orange |  |  | Green | Orange |
| Self | Green | \$4.00, 4.00 | \$2.00, 3.50 | Self | Green | \$4.00, 4.00 | \$4.50, 1.50 |
| | Orange | \$3.50, 2.00 | \$1.50, 1.50 | | Orange | \$1.50, 4.50 | \$2.00, 2.00 |

Table 6 presents results from Cox regressions that subsample participants according to their choices in these games. We find that primary results hold with statistical significance when sub-sampling only those subjects who played Green in Game C (Columns 3 and 4). Upon further subsampling (Columns 5 – 8), the magnitude of the effect is similar, but statistical significance fades with a dwindling sample size.

**Table 6:**
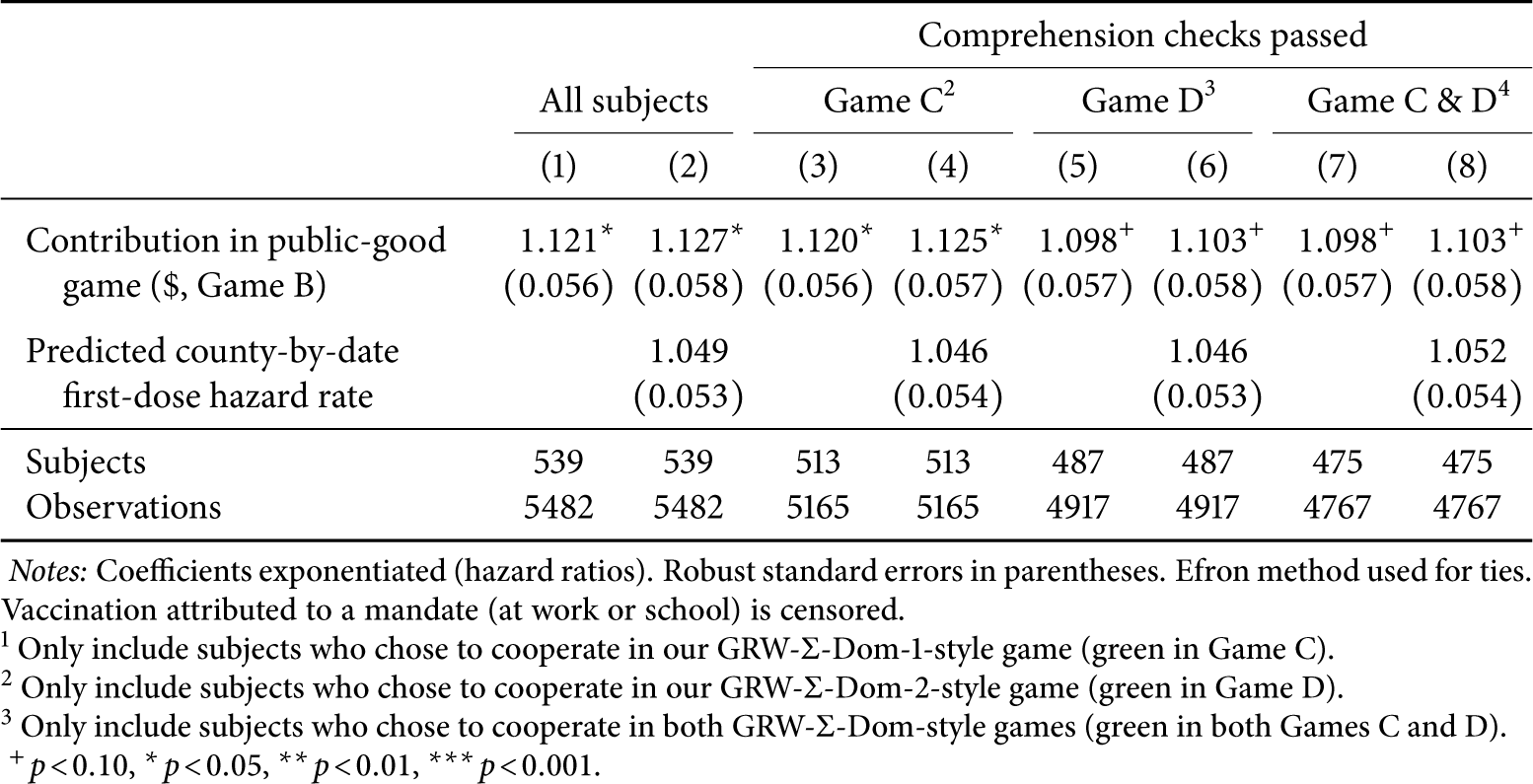
Cox regressions of vaccination hazard, conditional on comprehension checks.

### A.2. Additional regression analyses with individual covariates

Our primary analysis (as in Table 4) only included game behavior and the predicted vaccination hazard rate to preserve our degrees of freedom, considering our sample size. Here we offer additional Cox regressions that include individual co-variates collected in Session I.

First we include survey responses to questions about COVID-19 vaccination, shown in Table 7. As noted in Section 4.1, contribution in the public-good game is not significantly correlated with one’s response to the prompt “Getting vaccinated helps protect the health of my friends and family.” Perhaps care for one’s friends and family is relatively self-interested compared to contribution in a public-good game played online with strangers. Regardless, Table 7 shows that behavior in the public-good game is meaningful, even with all COVID-19 survey responses included (Column 2). Clearly this game is capturing social preferences that go beyond “protecting the health of family and friends” with COVID-19 vaccination. We caution that some of these regressors are collinear with contribution in the public-good game, possibly inflating standard errors and making individual coeffecients less meaningful.^22^

**Table 7:** Cox regressions of vaccination hazard with COVID-19 survey responses.

|  | (1) | (2) | (3) |
| --- | --- | --- | --- |
| Endowment to other participant (\$, Game A) | 1.020<br>(0.038) | | |
| Contribution in public-good game (\$, Game B) | 1.114<sup>+</sup><br>(0.065) | 1.119*<br>(0.064) | |
| Dislike shots or needles | 0.923<br>(0.050) | 0.922<br>(0.049) | 0.919<br>(0.052) |
| Difficult to find time to get vaccinated | 0.984<br>(0.063) | 0.985<br>(0.063) | 1.012<br>(0.065) |
| Worried about missing work to get vaccinated | 1.033<br>(0.082) | 1.036<br>(0.083) | 1.014<br>(0.082) |
| Worried about short-term side effects or allergic reactions | 0.887<sup>+</sup><br>(0.062) | 0.884<sup>+</sup><br>(0.062) | 0.884<sup>+</sup><br>(0.064) |
| Worried about long-term health risks of the vaccines | 0.834*<br>(0.064) | 0.836*<br>(0.065) | 0.837*<br>(0.064) |
| Wait for FDA approval of a vaccine | 0.882<br>(0.073) | 0.881<br>(0.073) | 0.873<sup>+</sup><br>(0.072) |
| Wait to see if it was safe for others | 1.162<br>(0.128) | 1.154<br>(0.127) | 1.162<br>(0.129) |
| Wait to see if it was effective for others | 0.894<br>(0.085) | 0.896<br>(0.086) | 0.885<br>(0.086) |
| Didn't/don't trust the vaccines | 1.082<br>(0.077) | 1.087<br>(0.078) | 1.087<br>(0.078) |
| Allows me to do more activities safely (socialize, travel, return to work or school) | 1.025<br>(0.126) | 1.024<br>(0.126) | 1.011<br>(0.128) |
| Getting vaccinated helps protect the health of my friends and family | 2.343***<br>(0.365) | 2.346***<br>(0.365) | 2.358***<br>(0.377) |
| A lottery, drawing, or other incentive did/would motivate me | 0.589***<br>(0.058) | 0.591***<br>(0.058) | 0.601***<br>(0.058) |
| Many of my friends and family are vaccinated | 1.164<sup>+</sup><br>(0.092) | 1.163<sup>+</sup><br>(0.091) | 1.170*<br>(0.091) |
| My friends or family want me to get vaccinated | 1.044<br>(0.080) | 1.050<br>(0.078) | 1.038<br>(0.077) |
| Pseudo-$R^2$ | 0.1465 | 0.1465 | 0.1446 |
| Subjects | 535 | 536 | 538 |
| Observations (subject-by-date) | 5426 | 5440 | 5468 |

Notes: Coefficients exponentiated (hazard ratios). Robust standard errors in parentheses. Efron method used for ties. Vaccination attributed to a mandate (at work or school) is censored. <sup>+</sup> $p < 0.10$ , \* $p < 0.05$ , \*\* $p < 0.01$ , \*\*\* $p < 0.001$ .

We also consider alternate definitions of a vaccination mandate. These are shown in Table 8. We consider vaccination attributed to a mandate for travel or social events, in addition to those for school or work (Columns 5 – 8). Recall that individuals who attribute their vaccination to a mandate are censored, so Columns 5 – 8 have more individuals censored (and thus have less power) than Columns 1 – 4. We also include specifications with a variety of individual co-variates (Columns 2, 4, 6, and 8), to which our results are largely robust. Naturally, these covariates do account for some variation explained by the games, thus slightly attenuating the coefficient associated with the public-good game. Despite the additional covariates and censoring (which reduces statistical power), our coefficient of interest remains similar.

**Table 8:**
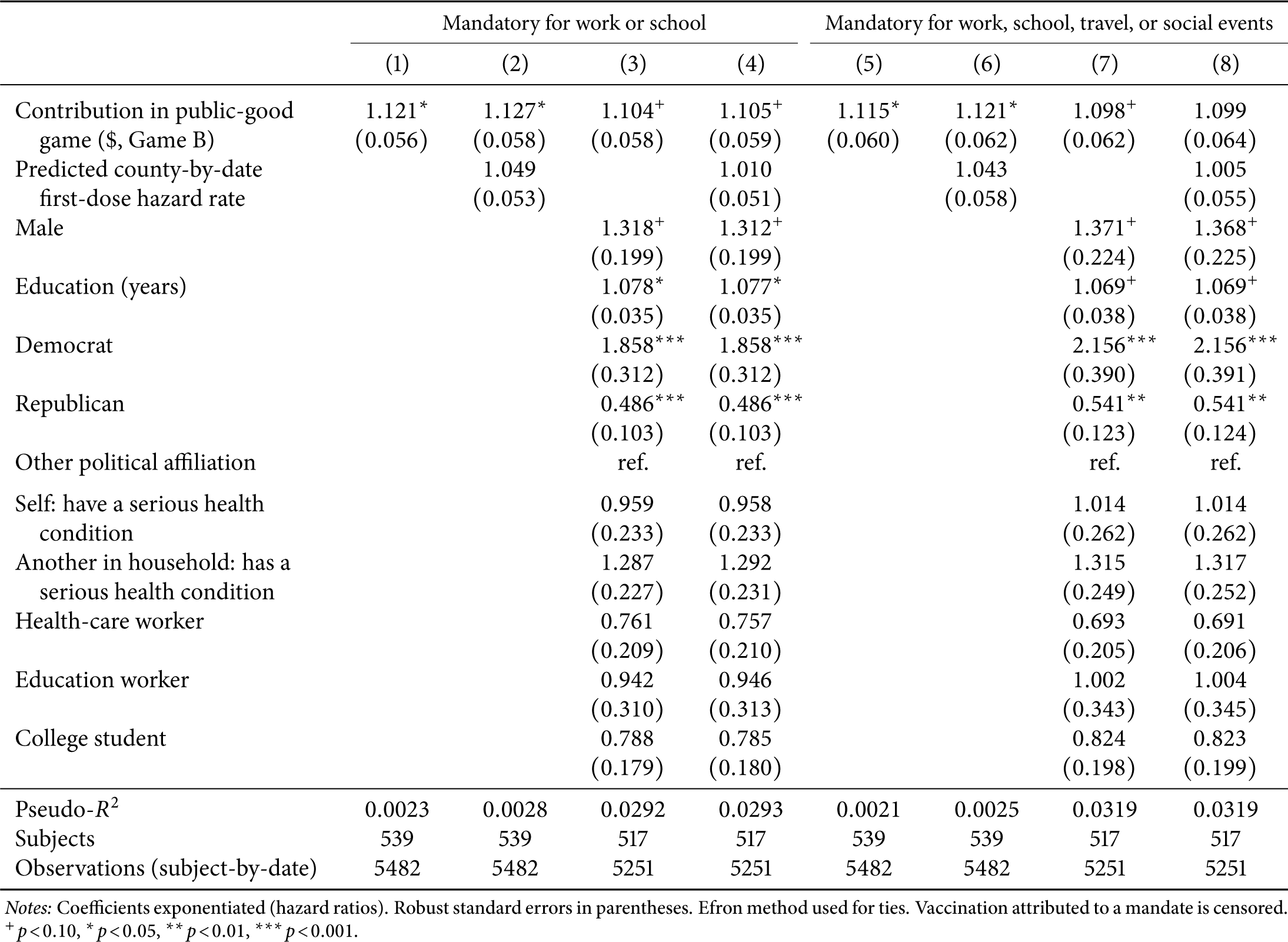
Additional Cox regressions of vaccination hazard (not attributed to a mandate)

## Appendix B. Robustness checks

### B.1. Standard errors

We report a variety of other standard errors for the Cox regressions in Table 9, finding no significant differences.

**Table 9:** Standard error comparison of Cox regressions of vaccination hazard.

|  | Huber-White |  | Cluster on U.S. state |  | Jackknife |  |
| --- | --- | --- | --- | --- | --- | --- |
|  | (1) | (2) | (3) | (4) | (5) | (6) |
| Contribution in public-good game (\$, Game B) | 1.121*<br>(0.056) | 1.127*<br>(0.058) | 1.121*<br>(0.058) | 1.127*<br>(0.059) | 1.121*<br>(0.056) | 1.127*<br>(0.058) |
| Predicted county-by-date first-dose hazard rate |  | 1.049<br>(0.053) |  | 1.049<br>(0.066) |  | 1.049<br>(0.054) |
| Subjects | 539 | 539 | 539 | 539 | 539 | 539 |
| Observations (subject-by-date) | 5482 | 5482 | 5482 | 5482 | 5482 | 5482 |

Notes: Coefficients exponentiated (hazard ratios). Robust standard errors in parentheses. Efron method used for ties. Vaccination attributed to a mandate (at work or school) is censored. <sup>+</sup> $p < 0.10$ , <sup>\*</sup> $p < 0.05$ , <sup>\*\*</sup> $p < 0.01$ , <sup>\*\*\*</sup> $p < 0.001$ .

### B.2. Schoenfeld residuals

Our Cox model assumes that hazard proportionality is time-invariant. We test this by inspecting Schoenfeld residuals for Column 4 of Table 4, shown in Figure 4. We find no significant effect of time on hazard proportionality (*χ*^2^ = 0.66, *p* = 0.72).

**Figure 4:**
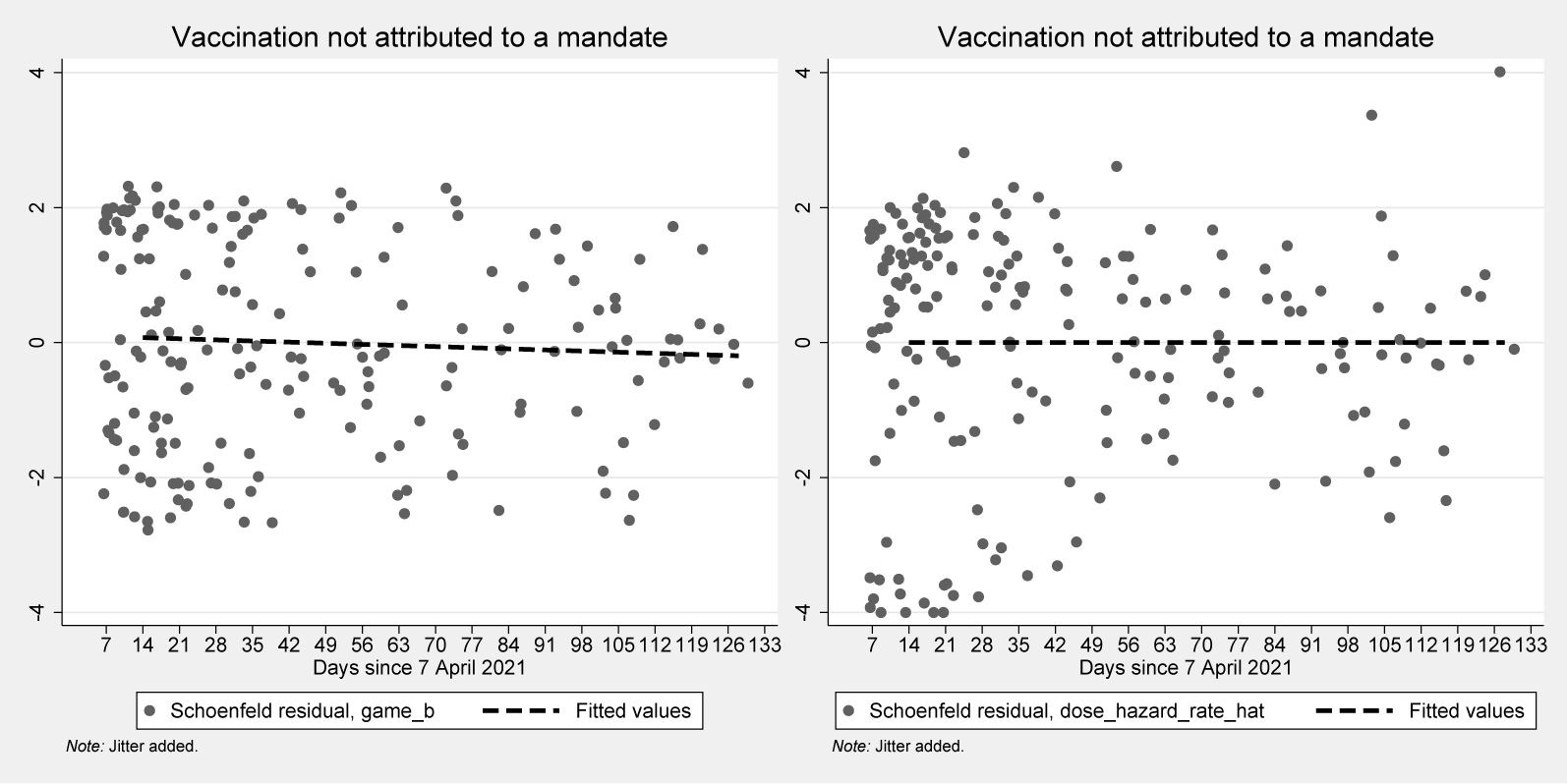
Schoenfeld residuals for the Cox regressions.

Analysis of Schoenfeld residuals test the proportional hazards assumption. We test whether proportionality of hazards varies with time by inspecting Schoenfeld residuals for Column 2 of Table 4, shown in Figure 4. We find no significant effect of time on hazard proportionality (*χ*^2^ = 0.66, *p* = 0.72).

### B.3. Results using alternative models

The hazard model of vaccination is the preferred specification, as it asserts that pro-social preferences influence vaccination uptake at any point in the time range of our study. Here we present results from alternative models that verify the robustness of our findings.

First we present a Tobit regression with the date-of-first-dose being predicted by pro-social preferences. We find that an additional dollar contribution in the public-good game predicts vaccination 6 days earlier (Table 10, Column 1). Compared to an individual who contributes $0 in the game, one who contributes the maximum of $4 is predicted to vaccinate 25 days earlier.

**Table 10:** Tobit regressions of date of first dose (relative to 1 April 2021)

|  | (1) | (2) |
| --- | --- | --- |
| Contribution in public-good game (\$, Game B) | -6.27*<br>(3.05) | -5.91*<br>(2.61) |
| Vaccinated under mandate | -82.17***<br>(16.04) | -57.21***<br>(13.50) |
| Predicted county-by-date first-dose hazard rate |  | -23.61***<br>(2.25) |
| Constant | 164.69***<br>(8.69) | 219.48***<br>(10.26) |
| Pseudo-$R^2$ | 0.0095 | 0.0444 |
| Observations | 539 | 539 |

Notes: Robust standard errors in parentheses. Tobit model with left-truncation at dose date 20 and right-censoring at dose date 136. Dependent variable, date of first dose, measures days since 1 April 2021.
+ $p < 0.10$ , \* $p < 0.05$ , \*\* $p < 0.01$ , \*\*\* $p < 0.001$ .

Next we offer results from ordinary least squares regressions that test whether being vaccinated by arbitrary points in time is predictive of pro-social preferences. As shown in Table 11, we find that individuals who vaccinate before most points in time contribute more in the public-good game.

**Table 11:** OLS regressions using time-of-first-dose indicators.

|  | (1) | (2) | (3) | (4) | (5) | (6) | (7) | (8) | (9) | (10) | (11) |
| --- | --- | --- | --- | --- | --- | --- | --- | --- | --- | --- | --- |
| Vacc. dose by... |  |  |  |  |  |  |  |  |  |  |  |
| April 30 | 0.416* |  |  |  |  |  |  |  |  |  |  |
|  | (0.194) |  |  |  |  |  |  |  |  |  |  |
| April 30 | -1.200** |  |  |  |  |  |  |  |  |  |  |
| mandated | (0.410) |  |  |  |  |  |  |  |  |  |  |
| May 10 |  | 0.280 |  |  |  |  |  |  |  |  |  |
|  |  | (0.173) |  |  |  |  |  |  |  |  |  |
| May 10 |  | -0.690<sup>+</sup> |  |  |  |  |  |  |  |  |  |
| mandated |  | (0.405) |  |  |  |  |  |  |  |  |  |
| May 20 |  |  | 0.291<sup>+</sup> |  |  |  |  |  |  |  |  |
|  |  |  | (0.162) |  |  |  |  |  |  |  |  |
| May 20 |  |  | -0.902** |  |  |  |  |  |  |  |  |
| mandated |  |  | (0.318) |  |  |  |  |  |  |  |  |
| May 31 |  |  |  | 0.385* |  |  |  |  |  |  |  |
|  |  |  |  | (0.154) |  |  |  |  |  |  |  |
| May 31 |  |  |  | -0.867** |  |  |  |  |  |  |  |
| mandated |  |  |  | (0.313) |  |  |  |  |  |  |  |
| June 10 |  |  |  |  | 0.342* |  |  |  |  |  |  |
|  |  |  |  |  | (0.148) |  |  |  |  |  |  |
| June 10 |  |  |  |  | -0.741* |  |  |  |  |  |  |
| mandated |  |  |  |  | (0.310) |  |  |  |  |  |  |
| June 20 |  |  |  |  |  | 0.265<sup>+</sup> |  |  |  |  |  |
|  |  |  |  |  |  | (0.146) |  |  |  |  |  |
| June 20 |  |  |  |  |  | -0.841** |  |  |  |  |  |
| mandated |  |  |  |  |  | (0.280) |  |  |  |  |  |
| June 30 |  |  |  |  |  |  | 0.257<sup>+</sup> |  |  |  |  |
|  |  |  |  |  |  |  | (0.144) |  |  |  |  |
| June 30 |  |  |  |  |  |  | -0.806** |  |  |  |  |
| mandated |  |  |  |  |  |  | (0.271) |  |  |  |  |
| July 10 |  |  |  |  |  |  |  | 0.282* |  |  |  |
|  |  |  |  |  |  |  |  | (0.141) |  |  |  |
| July 10 |  |  |  |  |  |  |  | -0.820** |  |  |  |
| mandated |  |  |  |  |  |  |  | (0.269) |  |  |  |
| July 20 |  |  |  |  |  |  |  |  | 0.298* |  |  |
|  |  |  |  |  |  |  |  |  | (0.140) |  |  |
| July 20 |  |  |  |  |  |  |  |  | -0.723** |  |  |
| mandated |  |  |  |  |  |  |  |  | (0.278) |  |  |
| July 31 |  |  |  |  |  |  |  |  |  | 0.249<sup>+</sup> |  |
|  |  |  |  |  |  |  |  |  |  | (0.137) |  |
| July 31 |  |  |  |  |  |  |  |  |  | -0.668** |  |
| mandated |  |  |  |  |  |  |  |  |  | (0.253) |  |
| Aug. 10 |  |  |  |  |  |  |  |  |  |  | 0.273* |
|  |  |  |  |  |  |  |  |  |  |  | (0.137) |
| Aug. 10 |  |  |  |  |  |  |  |  |  |  | -0.645** |
| mandated |  |  |  |  |  |  |  |  |  |  | (0.223) |
| Constant | 1.951*** | 1.955*** | 1.954*** | 1.922*** | 1.924*** | 1.950*** | 1.949*** | 1.938*** | 1.925*** | 1.937*** | 1.928*** |
|  | (0.068) | (0.070) | (0.072) | (0.074) | (0.075) | (0.077) | (0.078) | (0.079) | (0.080) | (0.083) | (0.085) |
| R<sup>2</sup> | 0.0130 | 0.0074 | 0.0126 | 0.0169 | 0.0142 | 0.0147 | 0.0142 | 0.0155 | 0.0144 | 0.0122 | 0.0139 |
| Observations | 539 | 539 | 539 | 539 | 539 | 539 | 539 | 539 | 539 | 539 | 539 |

Notes: Robust standard errors in parentheses. Dependent variable is contribution (\$) in the public-good game.
<sup>+</sup> $p < 0.10$ , \* $p < 0.05$ , \*\* $p < 0.01$ , \*\*\* $p < 0.001$ .

## Appendix C. Competing hazards

Because vaccination attributed to a mandate and vaccination not attributed to a mandate are likely informative of each other, we also consider the subhazard distribution model of competing hazards from Fine and Gray (1999) (see also Kalfleisch and Prentice 2002). With this model, once an individual experiences the competing event, she is not entirely removed from the risk set for the primary event of interest. For example, if an individual was not mandated by an employer to become vaccinated, they may have eventually chosen to do so on their own accord. Then removing them entirely from the risk set of non-mandated vaccination will bias our estimates. Note that we may analyze non-mandated vaccination as the primary event of interest with mandated vaccination as the competing event; we may also analyze mandated vaccination with non-mandated vaccination competing.

For each competing event type *k* ∈ {1, 2}—non-mandated and mandated vaccination—we model the hazard of the corresponding subdistribution as

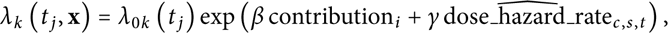

where *λ*_0*k*_ (*t _j_*) is the baseline hazard of the sub-distribution of event type *k*. This estimation is similar to the Cox model, except that people who experience a competing event are underweighted instead of being removed entirely from the risk set.

Results from the Cox and competing hazards regressions can jointly reveal how counterfactual events may bias our estimates. For example, suppose that greater contribution in the public-good game is associated with greater voluntary first-dose hazard and also with greater mandated firstdose hazard. When we observe an individual with a high public-good-game contribution who becomes vaccinated without a mandate, we cannot observe what she would have done had she waited to vaccinate and then been given a mandate. Because the competing event (mandated vaccination) also has a positive association with contribution, we would underestimate the effect of pro-social preferences on mandated vaccination. We would also simultaneously underestimate the effect of pro-social preferences on mandated vaccination. These relationships depend on whether the effect of pro-social preferences has the same or the opposing direction regarding the risk of each event type. However, note that because we do not observe counterfactuals, we cannot completely eliminate bias.

### C.1. Results

Columns 1 and 2 of Table 12 present results for vaccination not attributed to a mandate, treating vaccination attributed to a mandate as a competing event (and thus underweighting it). Likewise, Columns 3 and 4 contain results on mandated vaccination with non-mandated vaccination competing. Figure 5 depicts the results for both types of events. An additional dollar contribution in the public-good game is associated with 12% greater daily hazard of vaccination not attributed to a mandate. The regression for vaccination attributed to a mandate is slightly underpowered due to the additional censoring; our intent was to study voluntary vaccination.

**Figure 5:**
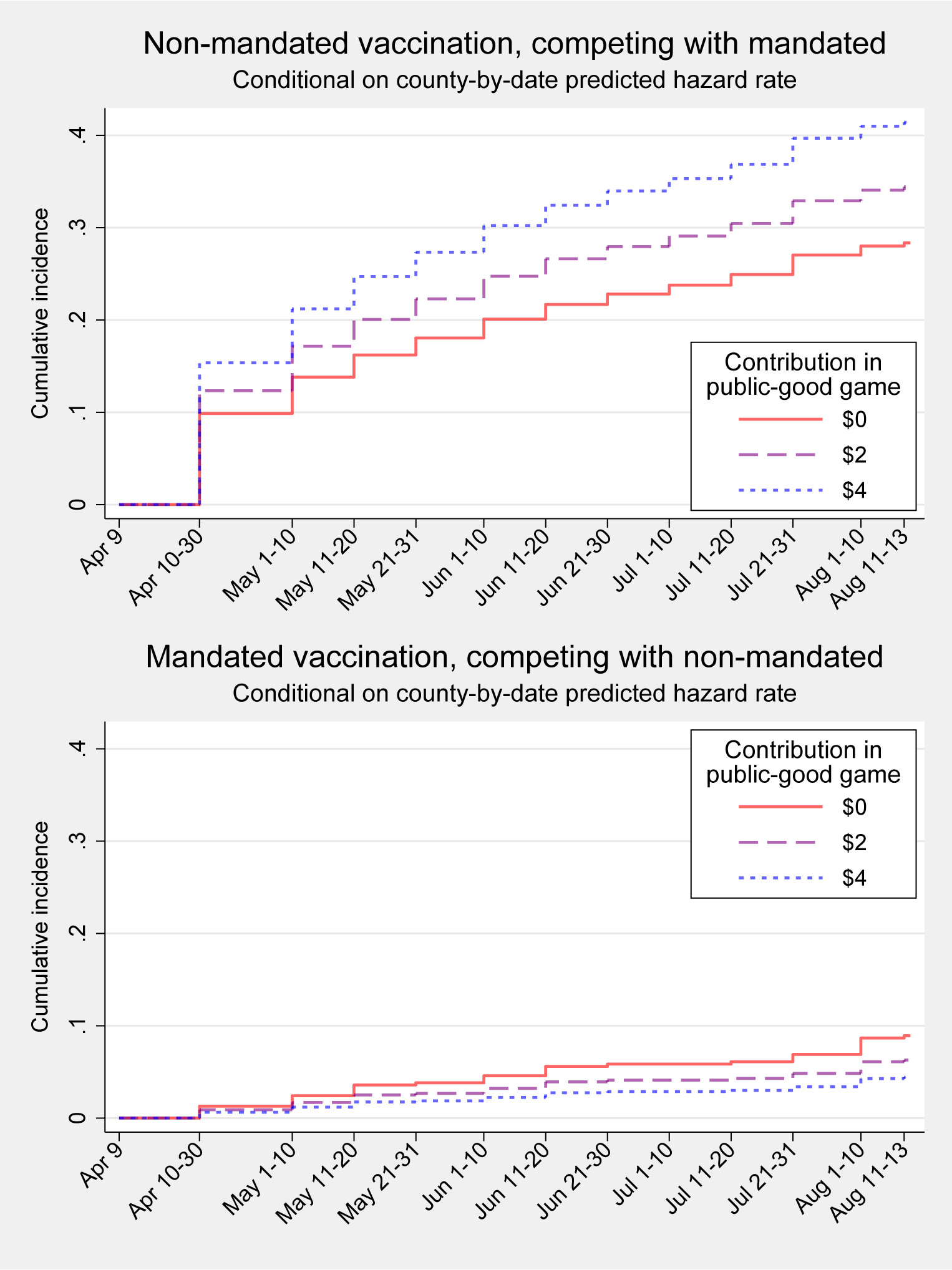
Regressions of competing vaccination events, non-mandated and mandated.

**Table 12:** Competing vaccination hazards regressions.

|  | Not due to a mandate<sup>1</sup> |  | Mandated<sup>2</sup> |  |
| --- | --- | --- | --- | --- |
|  | (1) | (2) | (3) | (4) |
| Contribution in public-good game (\$, Game B) | 1.122*<br>(0.054) | 1.125*<br>(0.055) | 0.832+<br>(0.082) | 0.834+<br>(0.083) |
| Predicted county-by-date first-dose hazard rate |  | 1.026<br>(0.047) |  | 1.106*<br>(0.055) |
| Subjects | 539 | 539 | 539 | 539 |
| Observations (subject-by-date) | 5482 | 5482 | 5482 | 5482 |

Notes: Coefficients exponentiated (hazard ratios). Robust standard errors in parentheses. Efron method used for ties.
<sup>1</sup> Vaccination attributed to a mandate (at work or school) is competing. <sup>2</sup> Vaccination not attributed to a mandate (at work or school) is competing. + $p < 0.10$ , \* $p < 0.05$ , \*\* $p < 0.01$ , \*\*\* $p < 0.001$ .

Comparing the Cox and competing hazards results, we find that the Cox regressions are fairly unbiased from the censored individuals who vaccinate due to a mandate. Further, mandated vaccination generally lags non-mandated vaccination, as shown in Figure 5. As a result, censoring mandated vaccination has little impact on the Cox regression. We conclude that an additional dollar of public-good contribution is associated with 1.12 times higher hazard of non-mandated vaccination. Therefore, an individual who contributed the maximal amount of $4 has a 58% greater hazard of voluntary vaccination relative to an individual who contributed nothing.

## Footnotes

5 Many of these people had already foregone the opportunity to vaccinate voluntarily, so perhaps had less concern for others.

6 Campos-Mercade et al. (2021b) used similar techniques to study COVID-19 prophylactic behavior.

7 Lab-in-the-field studies in the economics literature use similar methods (Gneezy and Imas 2017).

8 For the sake of comparability, we sought a sample of unvaccinated individuals once vaccination was widely available in the United States (the CDC removed eligibility considerations from its website on 19 April 2021). Thus our study naturally omits many of age 65 or with health problems who qualified for early vaccination. Individuals who obtained early vaccination may have been especially pro-social—or especially self-interested.

9 There are also at least a handful of other papers that consider pro-sociality or personality traits as predictors of vaccination. Zettler et al. (2022) finds COVID-19 health behaviors to be correlated with different personality traits such as agreeableness and honesty-humility. Bargain and Aminjonov (2020) examine political trust in Europe in relation to the pandemic, concluding dramatically that “degraded trust presents a risk for collective survival in the face of a pandemic”. Müller and Rau (2021) report survey data suggesting that risk tolerance negatively affects compliance to social distancing. Campos-Mercade et al. (2021b) correlate pro-sociality with health behavior in two surveys involving a broadly representative sample of the Swedish population, finding a relationship between stated prophylactic COVID-19 behaviors (e.g., wearing a face mask, reporting infection) and the willingness to expose another person to financial risk for one’s own financial benefit.

10 For example, Meier and Sprenger (2008) note that people who display more present bias in the lab also are more likely to carry large credit-card balances. Carpenter and Seki (2005) observe that conditional cooperation in a public-good game predicts group fishing productivity in Japan. Fehr and Leibbrandt (2008) show that public-good-game contributions and patience predict limits on common-pool resource extraction by Brazilian fishermen, and Barr and Zeitlin (2010) find that dictator-game allocations made by Ugandan teachers correlate with their actual (discretionary) teaching time. Carpenter and Myers (2010) find that dictator-game allocations by Vermont residents predict their willingness to volunteer to fight fires. Interestingly, Adena and Harke (2022) find that when a prospective donor is given an allusion to the COVID-19 pandemic, the donor makes a more generous contribution to the charity.

11 The completion of a two-dose vaccine induces greater time costs than completion of a single-dose vaccine, for example. Further, because a single-dose vaccination can be completed weeks earlier than a two-dose series by nature, the timing of a completed series is would be a poor comparison.

12 Our pre-analysis plan (Reddinger, Charness, and Levine 2021) specifies that we use games with a $4 endowment to measure social preferences; these two games fit this description. Because we are mainly interested in behavior in these games, we administer them first to minimize contamination from framing effects. Our pre-registration further specifies that we use two additional games as comprehension checks; for this we use two games styled after the “sigma-dominated” games that Gupta, Rigotti, and Wilson (2021) use as attention checks. Last, two games are appended for a separate study (these games were not mentioned in the pre-analysis plan). Participants thus play six games in total during Session II; Reddinger, Charness, and Levine (2023) provide survey instruments.

13 Philipson (1996) is an excellent example of using survival analysis to model the timing of MMR vaccination.

14 While our primary results only include mandates at school or work, Appendix A.2 offers similar results that also censor individuals who vaccinate due to a mandate for travel, social events, or restaurants.

15 The baseline hazard *h*_0_ (*t _j_*) is the probability that an individual gets vaccinated at time *t _j_*, conditional on the covariates being zero. (In some sense, this is analogous to the constant term in an OLS model.) When an individual contributes $1 instead of $0, the baseline hazard is multiplied by exp(*β*). The model thus assumes that at any point in time *t _j_*, the probability of vaccination is scaled proportionally by exp(*β*) (which is why the Cox model is also called the proportionalhazard model).

16 The University of California, Santa Barbara, Human Subjects Committee exempted our Protocol 60-20-0658. We obtained informed consent from all participants. We registered a pre-analysis plan with the American Economic Association as AEARCTR-0008216 (Reddinger, Charness, and Levine 2021). We use Stata 17 for analysis. Reddinger, Charness, and Levine (2023) provide data and source code.

17 All participants who completed Session I were invited to Session II, using the allow-list restriction (effectively an invitation list) on the Prolific platform.

18 The party breakdown of mandatory vaccination is interesting: 25.5% of vaccinated Republicans, but only 11.4% of Democrats and 14.5% of the remainder, said they vaccinated due to a mandate. We also ask unvaccinated respondents whether they would take the vaccine if it were mandatory, but we do not use these speculative responses in our analyses. With that caution in mind, when asked if they would comply with a vaccination mandate, 53.1% of Democrats responded “Yes”, while only 22.7% of Republicans and 26.6% of others stated they would comply.

19 Table 4 gives exponentiated coefficients, with exp(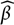) reported for the public-good game contribution, as used in our model in Section 3.2. Following the model, if the contribution is $0, then the baseline hazard is multiplied by exp(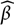×$0) = 1; if the contribution is $4, the baseline hazard is multiplied by (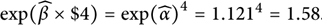.

20 We collapse the covariates into the lasso prediction to preserve degrees of freedom given our sample size. Moreover, the components are not individually meaningful in our context.

21 Campos-Mercade et al. (2021a) find that a $24-equivalent incentive increased vaccination by 4.2 p.p. (baseline 71.6%) in Sweden. Barber and West (2022) find that Ohio’s conditional cash lotteries increased the vaccination rate by 0.7 p.p. (baseline 46.5%). But Chang et al. (2021) find no effect of a $10 or $50 incentive among Medicaid plan members in California who delayed vaccination. Further, Serra-Garcia and Szech (2021) find that low monetary incentives ($10– $20) *decrease* vaccination, while high incentives ($100) increase vaccination. Clearly the success of monetary incentive programs depends on many factors, including the targeted population and the level of incentives.

22 See Online Appendix Table Z.1 for all correlation coefficients.

