## Supplementary material for "Vaccination as personal public-good provision": Online appendix (from another version of this preprint: version 3, 2024-04-15)

12 April 2024

#### Contents

1 Correlations of behavior in incentivized games with all survey data 1

2 Comparison of game choices by vaccination status as of study end-point 3

3 Session I survey instrument 4

4 Session II survey instrument 15

#### 1 Correlations of behavior in incentivized games with all survey data

Table Z.1 provides correlation coefficients between game choices and other data. Game A is our altruism game and Game B our public-good game. Game C and D are comprehension checks (see Appendix A.1 in the main text).

Table Z.1: Correlation coefficients between incentivized game behavior and all survey data

| | (1) Game A | (2) Game B | (3) Game C | (4) Game D |
|---|---|---|---|---|
| **Session I, Part I: Demographics** | | | | |
| Male | 0.123** | 0.044 | 0.044 | −0.017 |
| Age (years) | −0.126** | 0.044 | −0.028 | 0.005 |
| Education (years) | 0.003 | 0.052 | 0.042 | 0.095* |
| Democrat | −0.009 | 0.045 | 0.075+ | 0.046 |
| Republican | −0.024 | −0.043 | −0.077+ | 0.037 |
| Other political affiliation | ref. | ref. | ref. | ref. |
| Count in household: children under age 12 | −0.007 | −0.056 | 0.079+ | 0.062 |
| Count in household: children ages 12 to 17 | −0.092* | −0.067 | 0.023 | −0.030 |
| Count in household: adults ages 18 to 64 | 0.086* | 0.018 | 0.067 | −0.060 |
| Count in household: adults age 65 or over | 0.017 | 0.006 | −0.081+ | −0.089* |
| Self: have a serious health condition | −0.009 | −0.004 | −0.160*** | −0.033 |
| Another in household: has a serious health condition | −0.012 | 0.037 | −0.165*** | −0.014 |
| Income ($) | 0.019 | 0.089* | 0.116** | 0.018 |
| Assets ($) | 0.050 | 0.021 | 0.058 | 0.019 |
| College student | 0.074+ | −0.077+ | 0.040 | −0.029 |
| Health-care worker | −0.005 | 0.039 | 0.024 | −0.003 |
| Education worker | 0.026 | 0.073+ | 0.025 | 0.012 |

*Continued…*

Table Z.1: Correlation coefficients between incentivized game behavior and all survey data

| | (1) Game A | (2) Game B | (3) Game C | (4) Game D |
|---|---|---|---|---|
| **Session I, Part II: Vaccination history** | | | | |
| Have you received a dose of a COVID-19 vaccine? | 0.010 | 0.041 | 0.094$^{*}$ | 0.017 |
| Vaccinated not under mandate | 0.017 | 0.079$^{+}$ | 0.101$^{*}$ | 0.048 |
| Vaccinated under mandate | −0.012 | −0.070 | −0.008 | −0.059 |
| Date of first vaccine dose | −0.115$^{+}$ | −0.042 | −0.052 | −0.110$^{+}$ |
| **Session I, Part III: Vaccination attitudes[1]** | | | | |
| Dislike shots or needles | −0.050 | 0.023 | −0.009 | −0.020 |
| Difficult to find time to get vaccinated | 0.071$^{+}$ | 0.041 | 0.044 | −0.035 |
| Worried about missing work to get vaccinated | 0.039 | −0.042 | 0.017 | −0.042 |
| Worried about short-term side effects or allergic reactions | −0.084$^{+}$ | −0.113$^{**}$ | −0.086$^{*}$ | −0.025 |
| Worried about long-term health risks of the vaccines | −0.086$^{*}$ | −0.113$^{**}$ | −0.041 | 0.030 |
| Wait for FDA approval of a vaccine | −0.100$^{*}$ | −0.102$^{*}$ | −0.072$^{+}$ | −0.031 |
| Wait to see if it was safe for others | −0.102$^{*}$ | −0.117$^{**}$ | −0.082$^{+}$ | −0.003 |
| Wait to see if it was effective for others | −0.059 | −0.106$^{*}$ | −0.077$^{+}$ | −0.023 |
| Didn't/don't trust the vaccines | −0.066 | −0.097$^{*}$ | −0.050 | 0.038 |
| Mandatory for social events | 0.011 | 0.003 | −0.144$^{***}$ | −0.026 |
| Mandatory for travel | −0.040 | 0.001 | −0.093$^{*}$ | −0.063 |
| Mandatory for school | 0.072$^{+}$ | −0.013 | −0.031 | −0.088$^{*}$ |
| Mandatory for work | 0.011 | −0.019 | −0.007 | 0.023 |
| Allows me to do more activities safely (socialize, travel, return to work or school) | 0.001 | 0.063 | 0.007 | 0.001 |
| Getting vaccinated helps protect the health of my friends and family | 0.013 | 0.048 | 0.002 | −0.032 |
| A lottery, drawing, or other incentive did/would motivate me | −0.026 | 0.034 | −0.037 | 0.010 |
| Many of my friends and family are vaccinated | 0.009 | 0.057 | 0.028 | 0.027 |
| My friends or family want me to get vaccinated | −0.003 | −0.004 | −0.001 | −0.032 |
| Endowment to other participant ($, Game A) | 1.000 | 0.201$^{***}$ | | |
| **Session II, Part I: Incentivized games** | | | | |
| Contribution in public-good game ($, Game B) | 0.201$^{***}$ | 1.000 | 0.063 | 0.129$^{**}$ |
| GRW-style Sig-Dom 1, Game C | 0.118$^{**}$ | 0.063 | 1.000 | 0.334$^{***}$ |
| GRW-style Sig-Dom 2, Game D | 0.083$^{+}$ | 0.129$^{**}$ | 0.334$^{***}$ | 1.000 |
| **Session II, Part II: Pro-social attitudes[1]** | | | | |
| Helping others is usually a waste of time. | −0.073$^{+}$ | −0.203$^{***}$ | −0.031 | −0.095$^{*}$ |
| When given the opportunity, I enjoy aiding others who are in need. | 0.060 | 0.132$^{**}$ | 0.011 | −0.021 |
| Helping friends and family is one of the great joys in life. | 0.060 | 0.141$^{***}$ | 0.020 | 0.009 |
| It feels wonderful to assist others in need. | 0.039 | 0.150$^{***}$ | −0.006 | 0.009 |
| Volunteering to help someone is very rewarding. | 0.047 | 0.117$^{**}$ | −0.005 | −0.013 |
| I dislike giving directions to strangers who are lost. | −0.021 | −0.056 | 0.012 | −0.020 |
| Unless they are part of my family, helping the elderly isn't my responsibility. | 0.058 | −0.121$^{**}$ | 0.000 | 0.012 |
| If the person in front of me in the check-out line at a store was a few cents short, I would pay the difference. | 0.090$^{*}$ | 0.193$^{***}$ | 0.068 | 0.070 |
| I plan to donate my organs when I die with the hope that they will help someone else live. | 0.081$^{+}$ | 0.187$^{***}$ | 0.035 | 0.093$^{*}$ |
| Helping people does more harm than good because they come to rely on others and not themselves. | −0.137$^{**}$ | −0.182$^{***}$ | −0.088$^{*}$ | −0.047 |
| Observations | 549 | 549 | 549 | 549 |

$^{+}p < 0.10$, $^{*}p < 0.05$, $^{**}p < 0.01$, $^{***}p < 0.001$ [1] Likert scale of 5 "Strongly agree," 4 "Somewhat agree," 3 "Neither agree nor disagree," 2 "Somewhat disagree," and 1 "Strongly disagree."

#### 2 Comparison of game choices by vaccination status as of study end-point

The Kaplan-Meier curves shown in Figure 2 (in the main text) show the importance of the timing of vaccination without making any modeling assumptions. We can ignore vaccination timing and focus solely on vaccination status when the survey was conducted, as shown in Figure Z.1. These histograms give a snapshot of vaccination status as of 12–13 August 2021, thereby masking heterogeneity in the timing of vaccination. We thus also offer an alternative plot (Figure Z.2) that retains information on vaccination timing but is more difficult to interpret.

Figure Z.1: Histograms of game behavior by vaccination status as of August 2021

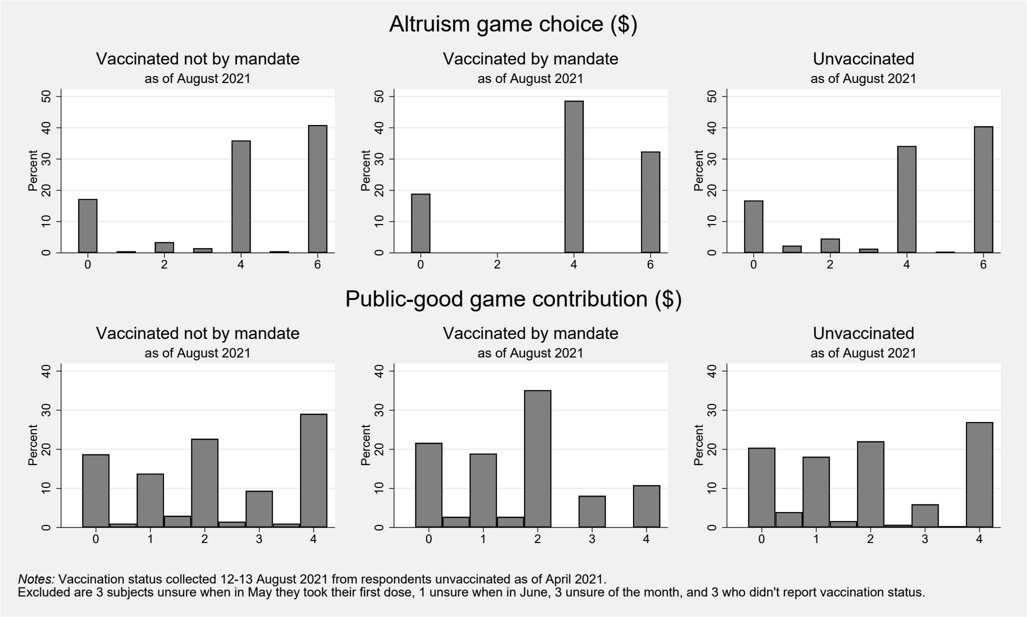


Figure Z.2: Participant counts by public-good-game contribution, date of first vaccine dose, and vaccination cause

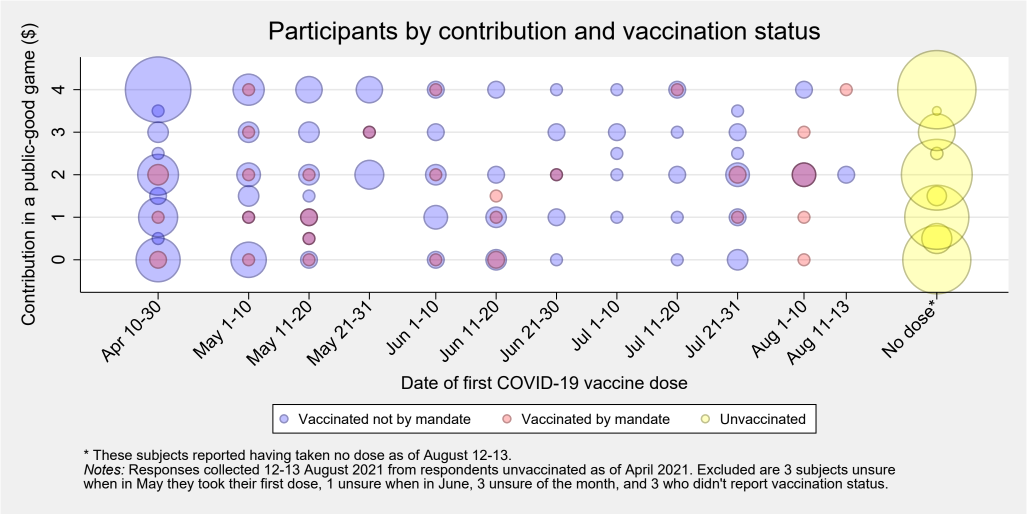

### 3 Session I survey instrument

#### 3.1 Logical flow

1. Show Block: Introduction
2. If Consent is not 1 then
   - End Survey
3. Show Block: DemogA
4. If Age is null or Gender is null then
   - End Survey
5. Show Block: DemogB
6. Show Block: VaxHistory
7. Set FirstDoseName to "first dose"
8. If VaxType is 1 then
   - Set FirstDoseName to "vaccine"
9. If VaxDose is 1 then
   - Show Block: VaxDate
10. If VaxDose is 1 then
    a) Show Block: VaxDose1Part1
    b) Randomizer: Evenly Present Elements
       - Show Block: VaxDose1Part2a
       - Show Block: VaxDose1Part2b
11. If VaxDose is not 1 then
    a) Show Block: VaxDose0Part1
    b) Randomizer: Evenly Present Elements
       - Show Block: VaxDose0Part2a
       - Show Block: VaxDose0Part2b
12. End Survey

#### 3.2 Element blocks

##### 3.2.1 Introduction

[Intro] **Survey about COVID-19 vaccination**

15 multiple-choice questions about your reasons for taking or not taking a COVID-19 vaccine.

15 multiple-choice demographic questions.

Estimated 5 minutes to complete. $1.00 payment.

This survey is completely anonymous.

[Consent] **Consent**

This is an academic research project to study what motivates people to obtain COVID-19 vaccination.

**The data we collect is anonymous and will not be linked to your identity in any way.**

You may choose to quit at any time. Risks are comparable to typical computer use. There is no direct benefit to you anticipated from your participation in this study.

If you have any questions about this research project, please contact Lucas Reddinger at.

If you have any questions regarding your rights and participation as a research subject, please contact the Human Subjects Committee at (805) 893-3807 or. Or write to the University of California, Human Subjects Committee, Office of Research, Santa Barbara, CA 93106-2050.

Participation in research is voluntary. Clicking the button labeled "I Consent" below will indicate that you have decided to participate as a research subject in the study described above.

- I CONSENT (1)
- I DO NOT CONSENT (0)

[Page Break]

###### 3.2.2 DemogA

[IntroDemog] **Please first answer this set of 12 demographic questions.**

[Age] How old are you?

- 18 to 29 (18)
- 30 to 49 (30)
- 50 to 64 (50)
- 65 or older (65)

[Gender] What is your gender?

- Male (2)
- Female (1)
- Other (0)

[Page Break]

###### 3.2.3 DemogB

[Educ] What is the highest grade (or year) of regular school you have completed?

- Less than High School (1)
- 9 (9)
- 10 (10)
- 11 (11)
- …
- 19 (19)
- 20+ (20)

[HHMembers] Consider everyone in your household, including yourself.

- How many are children under age 12? : ____________ (1)
- How many are children ages 12 to 17? : ____________ (2)
- How many are adults ages 18 to 64? : ____________ (3)
- How many are adults age 65 or over? : ____________ (4)
- Total : __[tabulated automatically]__

[Page Break]

[HealthSelf] Do you have a serious health condition such as high blood pressure, heart disease, lung disease, cancer, or diabetes?

- ○ Yes (1)
- ○ No (0)
- ○ Prefer not to say (9)

[HealthHH] Does someone else in your household have a serious health condition such as high blood pressure, heart disease, lung disease, cancer, or diabetes?

- ○ Yes (1)
- ○ No (0)
- ○ Prefer not to say (9)

[Page Break]

[USState] In which state do you currently reside?

- ○ Alabama (1)
- ○ …
- ○ I do not reside in the United States (53)

[Pols] Politically, which do you consider yourself as?

- ○ Republican (3)
- ○ Democrat (2)
- ○ Independent (1)
- ○ Other / None / No preference (0)
- ○ Don't know (9)

[Display This Question: If Pols is not 3 And Pols is not 2]

[PolsLean] Toward which party do you lean?

- ○ Republican (3)
- ○ Democrat (2)
- ○ Independent / Don't lean to either party (1)
- ○ Other party (0)
- ○ Don't know (9)

[Page Break]

[Income] Approximately what is your combined household income for the past 12 months? (Include all sources of income.)

- ○ Less than $10,000 (0)

- $10,000 to less than $20,000 (10)
- $20,000 to less than $30,000 (20)
- $30,000 to less than $40,000 (30)
- $40,000 to less than $50,000 (40)
- $50,000 to less than $60,000 (50)
- $60,000 or more (60)
- Don't know (999)

[Assets] Suppose your household needed money quickly, and your household cashed in all of its checking and savings accounts and investments. Approximately how much would this add up to?

- Less than $500 (0)
- $500 to less than $1,000 (0.5)
- $1,000 to less than $2,500 (1)
- $2,500 to less than $5,000 (2.5)
- $5,000 to less than $7,500 (5)
- $7,500 to less than $10,000 (7.5)
- $10,000 or more (10)
- Don't know (999)

[Page Break]

[College] Are you a student or employee at a college or university? (Select all that apply.)

- ☐ Undergraduate student (4)
- ☐ Graduate student (3)
- ☐ Other student (2)
- ☐ Employed at a college or university (1)
- ☐ No, neither (5)

[HCW] Do you or does someone in your household work in health care?

- Yes, myself (1)
- Yes, someone else in my household (2)
- Yes, both myself and someone else in my household (3)
- No (0)

[EdWorker] Do you or does someone in your household work at a school (any grade K-12)?

- Yes, myself (1)
- Yes, someone else in my household (2)
- Yes, both myself and someone else in my household (3)
- No (0)

[Page Break]

###### 3.2.4 VaxHistory

[VaxDose] Have you received a dose of a COVID-19 vaccine?

- Yes (1)

- No (0)
- Prefer not to say (9)

[Display This Question: If VaxDose is 1]

[VaxType] Did you receive a vaccine that requires two doses or one dose?

- Two doses (2)
- One dose (1)
- Don't know (0)
- Prefer not to say (9)

[Display This Question: If VaxType is 2]

[VaxIntent2] How likely are you to take the second dose?

- Already took both doses (7)
- Will definitely get the second dose (5)
- Will probably get the second dose (4)
- Unsure about getting the second dose (3)
- Will probably NOT get the second dose (2)
- Will definitely NOT get the second dose (1)
- Unsure / Don't know (0)
- Prefer not to say (9)

[Display This Question: If VaxDose is 0]

[VaxIntent1] How likely are you to get a COVID-19 vaccine?

- Will definitely get a COVID-19 vaccine (5)
- Will probably get a COVID-19 vaccine (4)
- Unsure about getting a COVID-19 vaccine (3)
- Will probably NOT get a COVID-19 vaccine (2)
- Will definitely NOT get a COVID-19 vaccine (1)
- Unsure / Don't know (0)
- Prefer not to say (9)

##### 3.2.5 VaxDate

[DoseMonth] When did you receive your [FirstDoseName]?

- April or earlier (4)
- May (5)
- June (6)
- July (7)
- August (8)
- Unsure / Don't know (99)

[Display This Question: If DoseMonth is 5 Or DoseMonth is 6 Or DoseMonth is 7 Or DoseMonth is 8]

[DoseTimeOfMonth] When in [DoseMonth]?

- Early in the month (Day 1 - 10) (1)
- Middle of the month (Day 11 - 20) (11)
- Late in the month (Day 21 - 31) (21)
- Unsure / Don't know (99)

###### 3.2.6 VaxDose1Part1

[Q1] *Think about the time before you were vaccinated for COVID-19. Did you delay vaccination for this reason?*

I don't like shots or needles.

- Strongly agree (5)
- Somewhat agree (4)
- Neither agree nor disagree (3)
- Somewhat disagree (2)
- Strongly disagree (1)

[Q2] *Think about the time before you were vaccinated for COVID-19. Did you delay vaccination for this reason?*

It was difficult to find time to get vaccinated.

- Strongly agree (5)
- Somewhat agree (4)
- Neither agree nor disagree (3)
- Somewhat disagree (2)
- Strongly disagree (1)

[Q3] *Think about the time before you were vaccinated for COVID-19. Did you delay vaccination for this reason?*

I was worried about to missing work to get vaccinated.

- Strongly agree (5)
- Somewhat agree (4)
- Neither agree nor disagree (3)
- Somewhat disagree (2)
- Strongly disagree (1)

[Q4] *Think about the time before you were vaccinated for COVID-19. Did you delay vaccination for this reason?*

I was worried about short-term side effects or an allergic reaction from a vaccine.

- Strongly agree (5)
- Somewhat agree (4)
- Neither agree nor disagree (3)
- Somewhat disagree (2)
- Strongly disagree (1)

[Q5] *Think about the time before you were vaccinated for COVID-19. Did you delay vaccination for this reason?*

I was worried about long-term health risks of the vaccines.

- ○ Strongly agree (5)
- ○ Somewhat agree (4)
- ○ Neither agree nor disagree (3)
- ○ Somewhat disagree (2)
- ○ Strongly disagree (1)

[Q6] *Think about the time before you were vaccinated for COVID-19. Did you delay vaccination for this reason?*

I wanted to wait for FDA approval of a vaccine.

- ○ Strongly agree (5)
- ○ Somewhat agree (4)
- ○ Neither agree nor disagree (3)
- ○ Somewhat disagree (2)
- ○ Strongly disagree (1)

[Q7] *Think about the time before you were vaccinated for COVID-19. Did you delay vaccination for this reason?*

I wanted to wait to see if it was safe for others.

- ○ Strongly agree (5)
- ○ Somewhat agree (4)
- ○ Neither agree nor disagree (3)
- ○ Somewhat disagree (2)
- ○ Strongly disagree (1)

[Q8] *Think about the time before you were vaccinated for COVID-19. Did you delay vaccination for this reason?*

I wanted to wait to see if it was effective for others.

- ○ Strongly agree (5)
- ○ Somewhat agree (4)
- ○ Neither agree nor disagree (3)
- ○ Somewhat disagree (2)
- ○ Strongly disagree (1)

[Q9] *Think about the time before you were vaccinated for COVID-19. Did you delay vaccination for this reason?*

I wanted to wait for FDA approval of a vaccine.

- ○ Strongly agree (5)
- ○ Somewhat agree (4)
- ○ Neither agree nor disagree (3)
- ○ Somewhat disagree (2)
- ○ Strongly disagree (1)

[Q10] Did you get vaccinated because it would be mandatory or required for you somehow? (Choose all that apply.)

- ☐ Yes, for work (1)
- ☐ Yes, for school (2)
- ☐ Yes, for travel (3)
- ☐ Yes, for social events or restaurants (4)
- ☐ No, none of these (5)

[Q11] Getting vaccinated allows me to do more activities safely (socialize, travel, return to work or school).

- Strongly agree (5)
- Somewhat agree (4)
- Neither agree nor disagree (3)
- Somewhat disagree (2)
- Strongly disagree (1)

[Q12] Getting vaccinated helps protect the health of my friends and family.

- Strongly agree (5)
- Somewhat agree (4)
- Neither agree nor disagree (3)
- Somewhat disagree (2)
- Strongly disagree (1)

[Q13] A lottery, drawing, or other incentive motivated me to get vaccinated.

- Strongly agree (5)
- Somewhat agree (4)
- Neither agree nor disagree (3)
- Somewhat disagree (2)
- Strongly disagree (1)

##### 3.2.7 VaxDose1Part2a

[Q14] Many of my friends and family are vaccinated.

- Strongly agree (5)
- Somewhat agree (4)
- Neither agree nor disagree (3)
- Somewhat disagree (2)
- Strongly disagree (1)

[Q15] My friends or family wanted me to get vaccinated.

- Strongly agree (5)
- Somewhat agree (4)
- Neither agree nor disagree (3)
- Somewhat disagree (2)
- Strongly disagree (1)

##### 3.2.8 VaxDose1Part2b

[Q16] Many of my friends and family are ***not*** vaccinated.

- Strongly agree (1)
- Somewhat agree (2)
- Neither agree nor disagree (3)

- Somewhat disagree (4)
- Strongly disagree (5)

[Q17] My friends or family wanted me to ***not*** get vaccinated.

- Strongly agree (1)
- Somewhat agree (2)
- Neither agree nor disagree (3)
- Somewhat disagree (4)
- Strongly disagree (5)

###### 3.2.9 VaxDose0Part1

[Q21] *Are you delaying or avoiding COVID-19 vaccination for this reason?*

I don't like shots or needles.

- Strongly agree (5)
- Somewhat agree (4)
- Neither agree nor disagree (3)
- Somewhat disagree (2)
- Strongly disagree (1)

[Q22] *Are you delaying or avoiding COVID-19 vaccination for this reason?*

It is difficult to find time to get vaccinated.

- Strongly agree (5)
- Somewhat agree (4)
- Neither agree nor disagree (3)
- Somewhat disagree (2)
- Strongly disagree (1)

[Q23] *Are you delaying or avoiding COVID-19 vaccination for this reason?*

I am worried about missing work to get vaccinated.

- Strongly agree (5)
- Somewhat agree (4)
- Neither agree nor disagree (3)
- Somewhat disagree (2)
- Strongly disagree (1)

[Q24] *Are you delaying or avoiding COVID-19 vaccination for this reason?*

I am worried about short-term side effects or an allergic reaction from a vaccine.

- Strongly agree (5)
- Somewhat agree (4)
- Neither agree nor disagree (3)
- Somewhat disagree (2)

- Strongly disagree (1)

[Q25] *Are you delaying or avoiding COVID-19 vaccination for this reason?*

I am worried about long-term health risks of the vaccines.

- Strongly agree (5)
- Somewhat agree (4)
- Neither agree nor disagree (3)
- Somewhat disagree (2)
- Strongly disagree (1)

[Q26] *Are you delaying or avoiding COVID-19 vaccination for this reason?*

I want to wait for FDA approval of a vaccine.

- Strongly agree (5)
- Somewhat agree (4)
- Neither agree nor disagree (3)
- Somewhat disagree (2)
- Strongly disagree (1)

[Q27] *Are you delaying or avoiding COVID-19 vaccination for this reason?*

I want to wait to see if it is safe for others.

- Strongly agree (5)
- Somewhat agree (4)
- Neither agree nor disagree (3)
- Somewhat disagree (2)
- Strongly disagree (1)

[Q28] *Are you delaying or avoiding COVID-19 vaccination for this reason?*

I want to wait to see if it is effective for others.

- Strongly agree (5)
- Somewhat agree (4)
- Neither agree nor disagree (3)
- Somewhat disagree (2)
- Strongly disagree (1)

[Q29] *Are you delaying or avoiding COVID-19 vaccination for this reason?*

I don't trust the vaccines.

- Strongly agree (5)
- Somewhat agree (4)
- Neither agree nor disagree (3)
- Somewhat disagree (2)
- Strongly disagree (1)

[Q30] Would you get vaccinated if it were mandatory or required for you somehow? (Choose all that apply.)

- ☐ Yes, for work (1)
- ☐ Yes, for school (2)
- ☐ Yes, for travel (3)
- ☐ Yes, for social events or restaurants (4)
- ☐ No, none of these (5)

[Q31] Getting vaccinated would allow me to do more activities safely (socialize, travel, return to work or school).

- ○ Strongly agree (5)
- ○ Somewhat agree (4)
- ○ Neither agree nor disagree (3)
- ○ Somewhat disagree (2)
- ○ Strongly disagree (1)

[Q32] Getting vaccinated would help protect the health of my friends and family.

- ○ Strongly agree (5)
- ○ Somewhat agree (4)
- ○ Neither agree nor disagree (3)
- ○ Somewhat disagree (2)
- ○ Strongly disagree (1)

[Q33] A lottery, drawing, or other incentive would motivate me get vaccinated.

- ○ Strongly agree (5)
- ○ Somewhat agree (4)
- ○ Neither agree nor disagree (3)
- ○ Somewhat disagree (2)
- ○ Strongly disagree (1)

###### 3.2.10 VaxDose0Part2a

[Q34] Many of my friends and family are vaccinated.

- ○ Strongly agree (5)
- ○ Somewhat agree (4)
- ○ Neither agree nor disagree (3)
- ○ Somewhat disagree (2)
- ○ Strongly disagree (1)

[Q35] My friends or family want me to get vaccinated.

- ○ Strongly agree (5)
- ○ Somewhat agree (4)
- ○ Neither agree nor disagree (3)
- ○ Somewhat disagree (2)
- ○ Strongly disagree (1)

###### 3.2.11 VaxDose0Part2b

[Q36] Many of my friends and family are ***not*** vaccinated.

- Strongly agree (1)
- Somewhat agree (2)
- Neither agree nor disagree (3)
- Somewhat disagree (4)
- Strongly disagree (5)

[Q37] My friends or family want me to ***not*** get vaccinated.

- Strongly agree (1)
- Somewhat agree (2)
- Neither agree nor disagree (3)
- Somewhat disagree (4)
- Strongly disagree (5)

### 4 Session II survey instrument

#### 4.1 Logical flow

1. Show Block: Introduction
2. If Consent is not 1 then
    - End Survey
3. Show Block: Instructions
4. Show Block: ScenarioA
5. Show Block: ScenarioB
6. Show Block: ScenarioCDEF
7. Show Block: SocialSurvey
8. End Survey

#### 4.2 Element blocks

##### 4.2.1 Introduction

[Intro] **Survey about decision-making**

6 decisions about monetary payments.

10 multiple-choice questions about social behavior.

Estimated 10 minutes to complete. $1.00 payment up-front.

You will likely earn a bonus of $4 or more!

[Consent] **Consent**

This is an academic research project to study how people make decisions.

The data we collect is anonymous and will not be linked to your identity in any way.

You may choose to quit at any time. Risks are comparable to typical computer use. There is no direct benefit to you anticipated from your participation in this study.

If you have any questions about this research project, please contact Gary Charness at.

If you have any questions regarding your rights and participation as a research subject, please contact the Human Subjects Committee at (805) 893-3807 or. Or write to the University of California, Human Subjects Committee, Office of Research, Santa Barbara, CA 93106-2050.

Participation in research is voluntary. Clicking the button labeled "I Consent" below will indicate that you have decided to participate as a research subject in the study described above.

- I CONSENT (1)
- I DO NOT CONSENT (0)

[Page Break]

###### 4.2.2 Instructions

[Instructions] This survey contains **scenarios** that can result in **substantial bonus payments**.

You will make one decision for each scenario.

After you submit your survey, the computer will randomly choose a scenario and you will be paid the appropriate bonus.

Bonus payments will be made within 24 hours.

[Page Break]

###### 4.2.3 ScenarioA

[ChoiceA] **Scenario A (1 of 6)**

In this scenario, you are matched with one survey participant. A computerized coin toss will determine what happens.

**If HEADS:**

You get a $4 bonus
The other participant gets a bonus amount that was chosen by you

**If TAILS:**

You get a bonus amount chosen by the other participant
The other participant gets a $4 bonus

**Choose what happens if the coin toss is HEADS:**

- The other person gets a $0 bonus. You get a $4 bonus. (0)
- The other person gets a $1 bonus. You get a $4 bonus. (1)
- The other person gets a $2 bonus. You get a $4 bonus. (2)
- The other person gets a $3 bonus. You get a $4 bonus. (3)
- The other person gets a $4 bonus. You get a $4 bonus. (4)
- The other person gets a $5 bonus. You get a $4 bonus. (5)
- The other person gets a $6 bonus. You get a $4 bonus. (6)

[Page Break]

###### 4.2.4 ScenarioB

**Scenario B (2 of 6)**

In this scenario you can earn up to a $10 bonus.

The amount depends on your decision and the decision of three other survey participants.

Please read the following instructions carefully!

[Page Break]

You are matched with three other survey participants

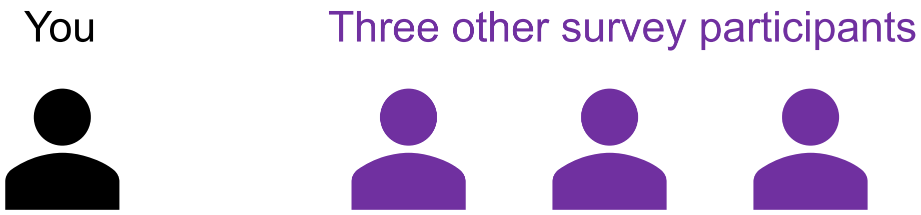


[Page Break]

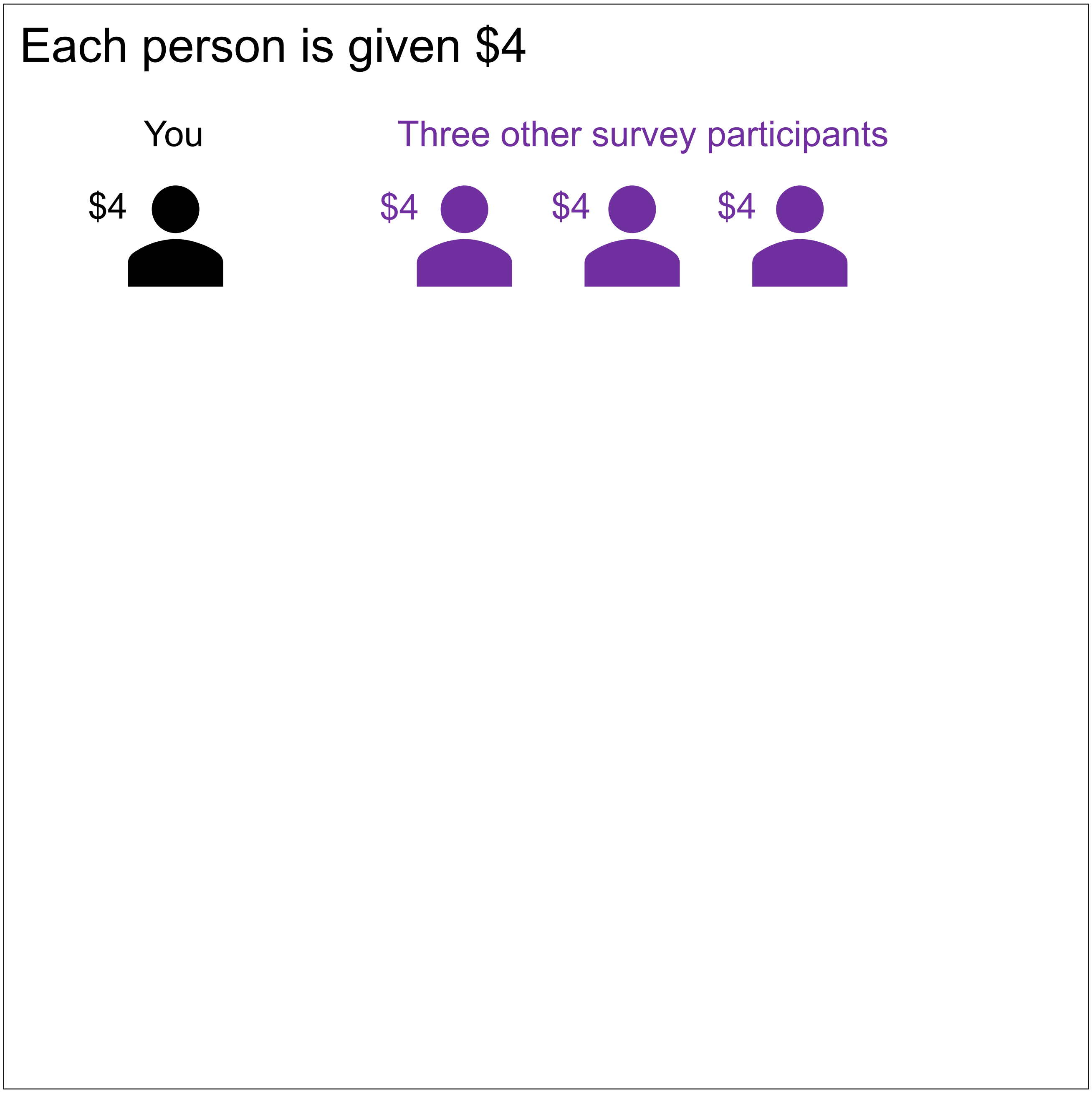
Each person is given $4
You
Three other survey participants
$4
$4
$4
$4

[Page Break]

#### Each person chooses how much to contribute to a pot

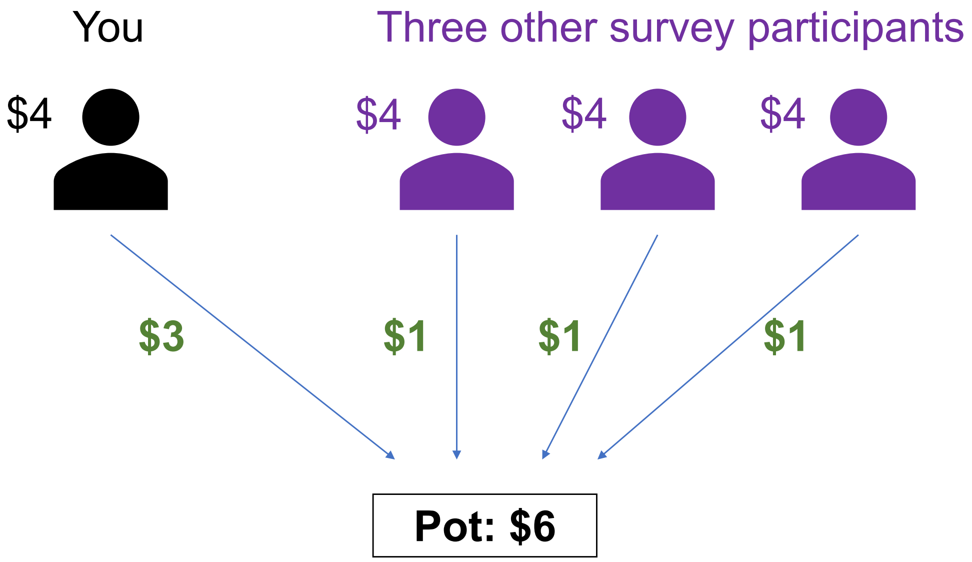


In this example, you choose to put in $3,
and every other person chooses to put in $1

[Page Break]

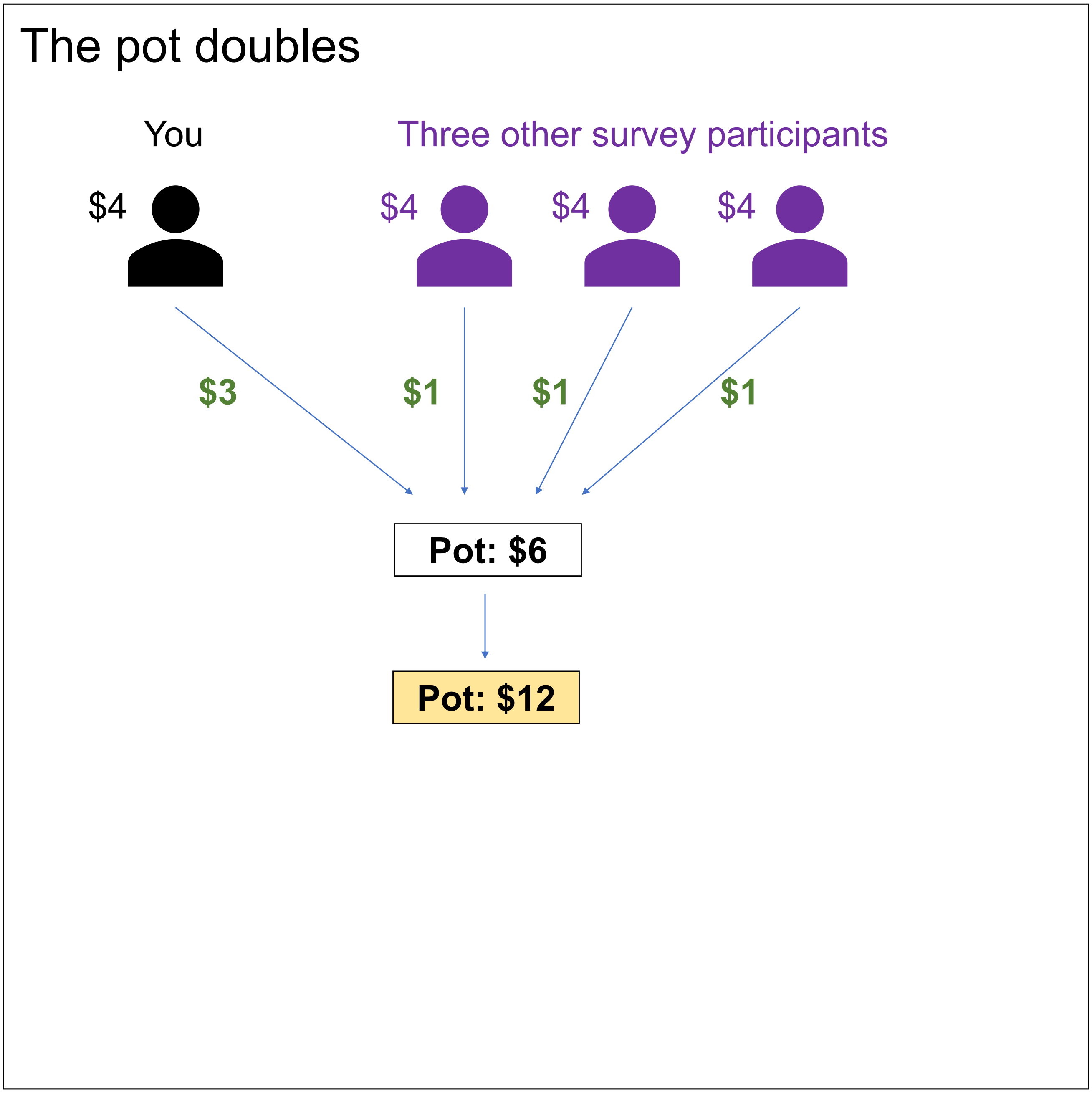
The pot doubles
You
Three other survey participants
$4
$4
$4
$4
$3
$1
$1
$1
Pot: $6
Pot: $12

[Page Break]

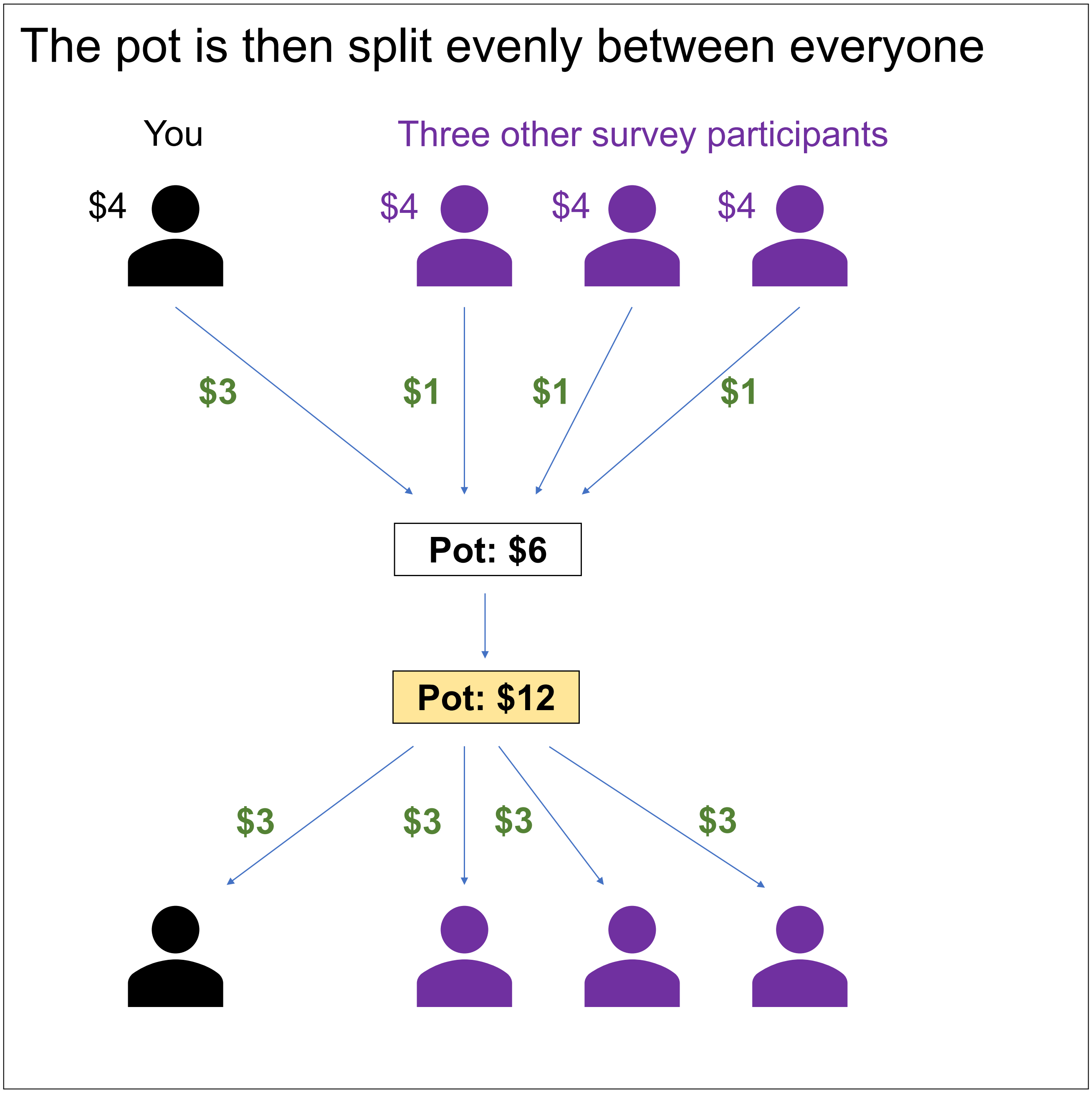
The pot is then split evenly between everyone
You
Three other survey participants
$4
$4
$4
$4
$3
$1
$1
$1
Pot: $6
Pot: $12
$3
$3
$3
$3

[Page Break]

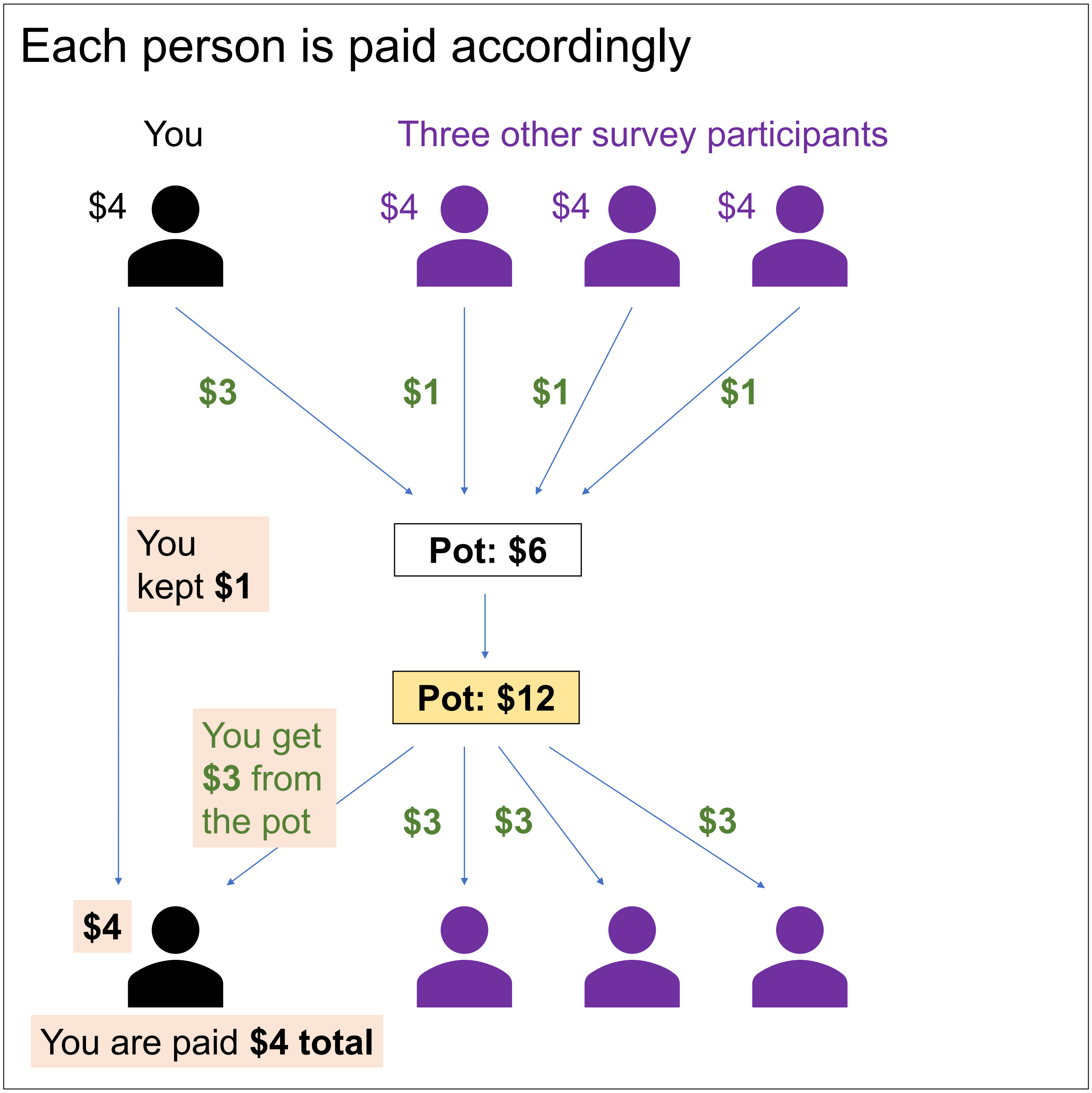
Each person is paid accordingly
You
Three other survey participants
$4
$4
$4
$4
$3
$1
$1
$1
You kept $1
Pot: $6
Pot: $12
You get $3 from the pot
$3
$3
$3
$4
You are paid $4 total

[Page Break]

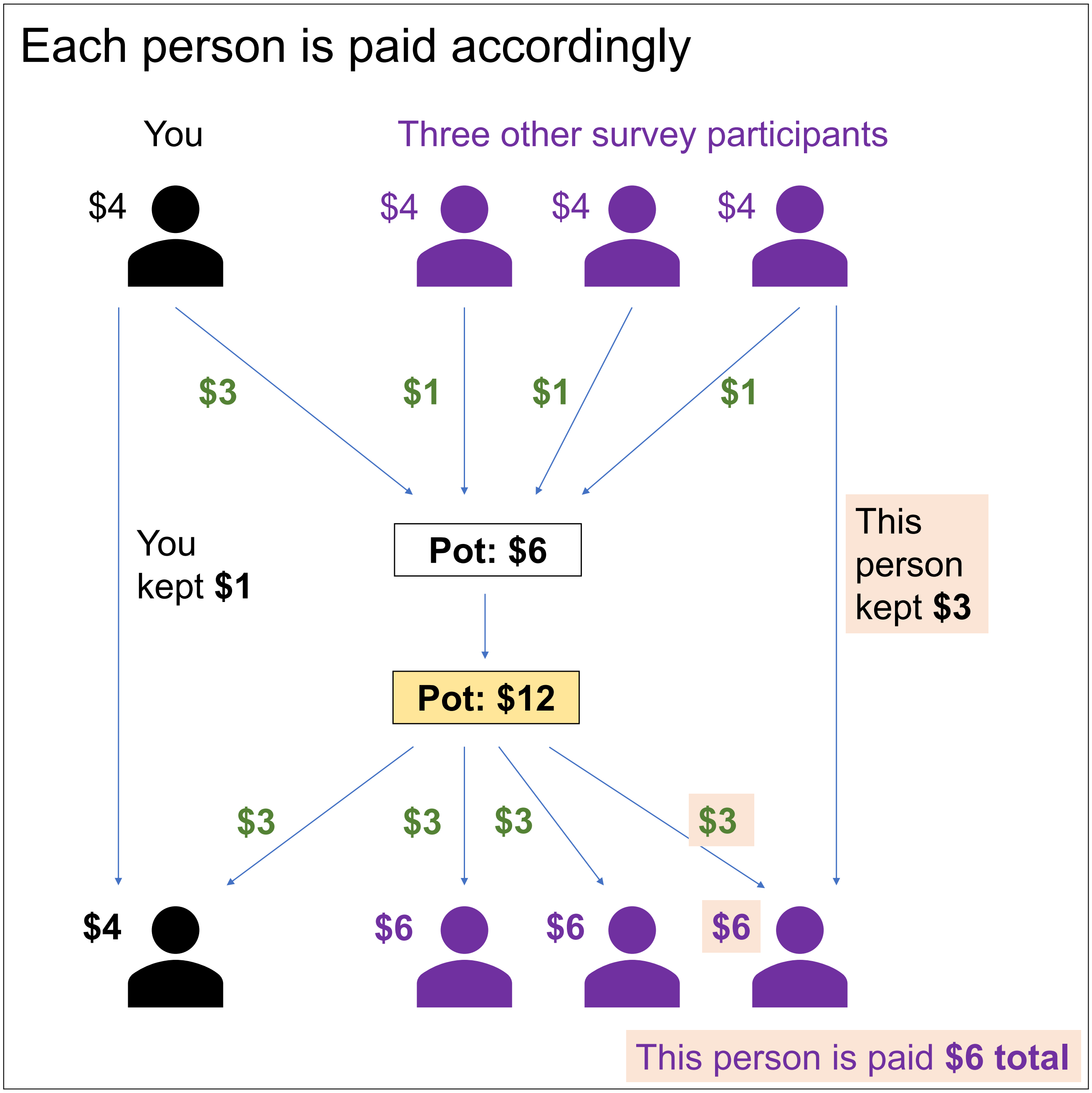
Each person is paid accordingly
You
Three other survey participants
$4
$4
$4
$4
$3
$1
$1
$1
You
kept $1
Pot: $6
This
person
kept $3
Pot: $12
$3
$3
$3
$3
$4
$6
$6
$6
This person is paid $6 total

[Page Break]

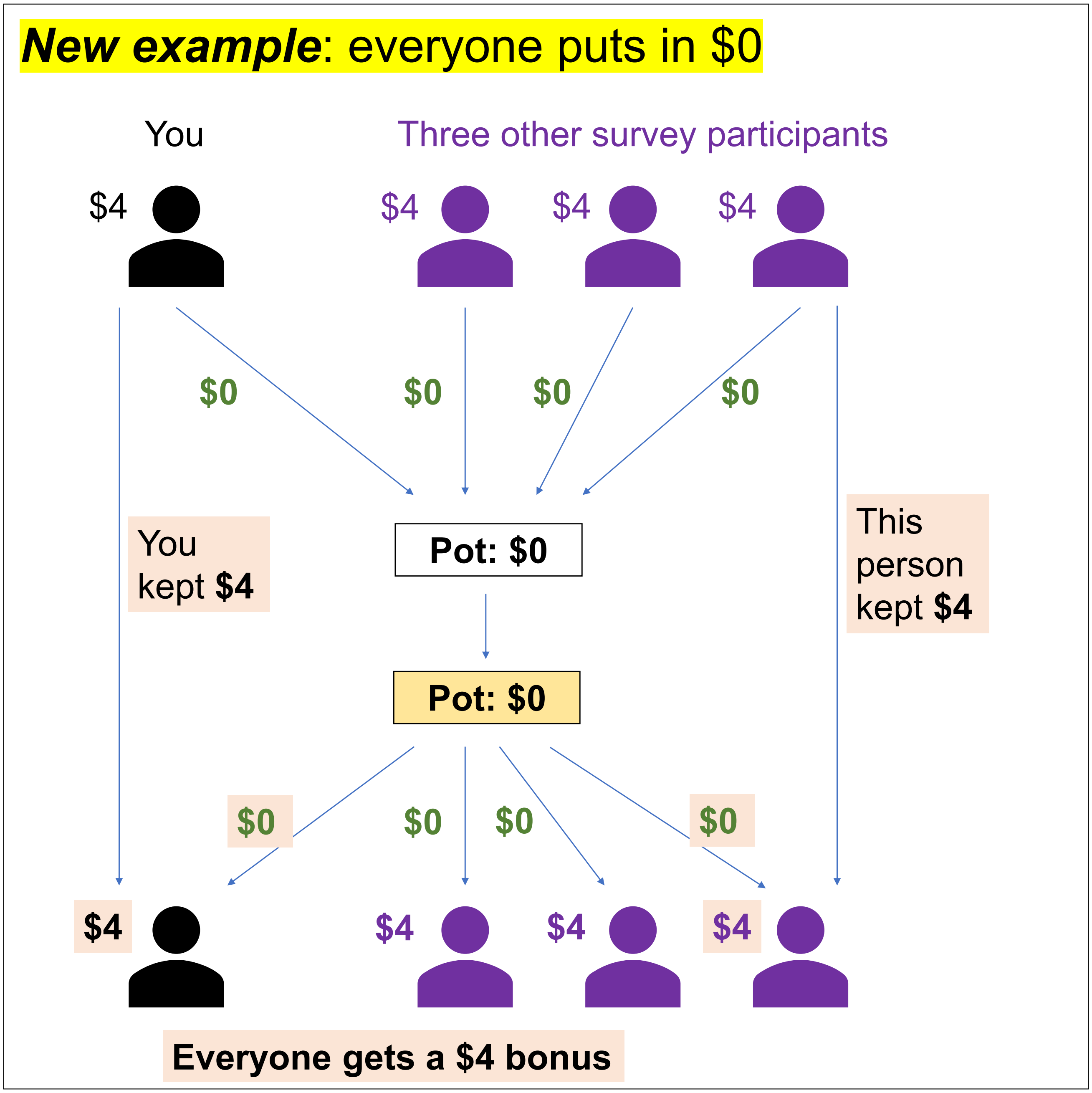
***New example***: everyone puts in $0
You
Three other survey participants
$4
$4
$4
$4
$0
$0
$0
$0
You kept **$4**
Pot: $0
This person kept **$4**
Pot: $0
$0
$0
$0
$0
$4
$4
$4
$4
Everyone gets a $4 bonus

[Page Break]

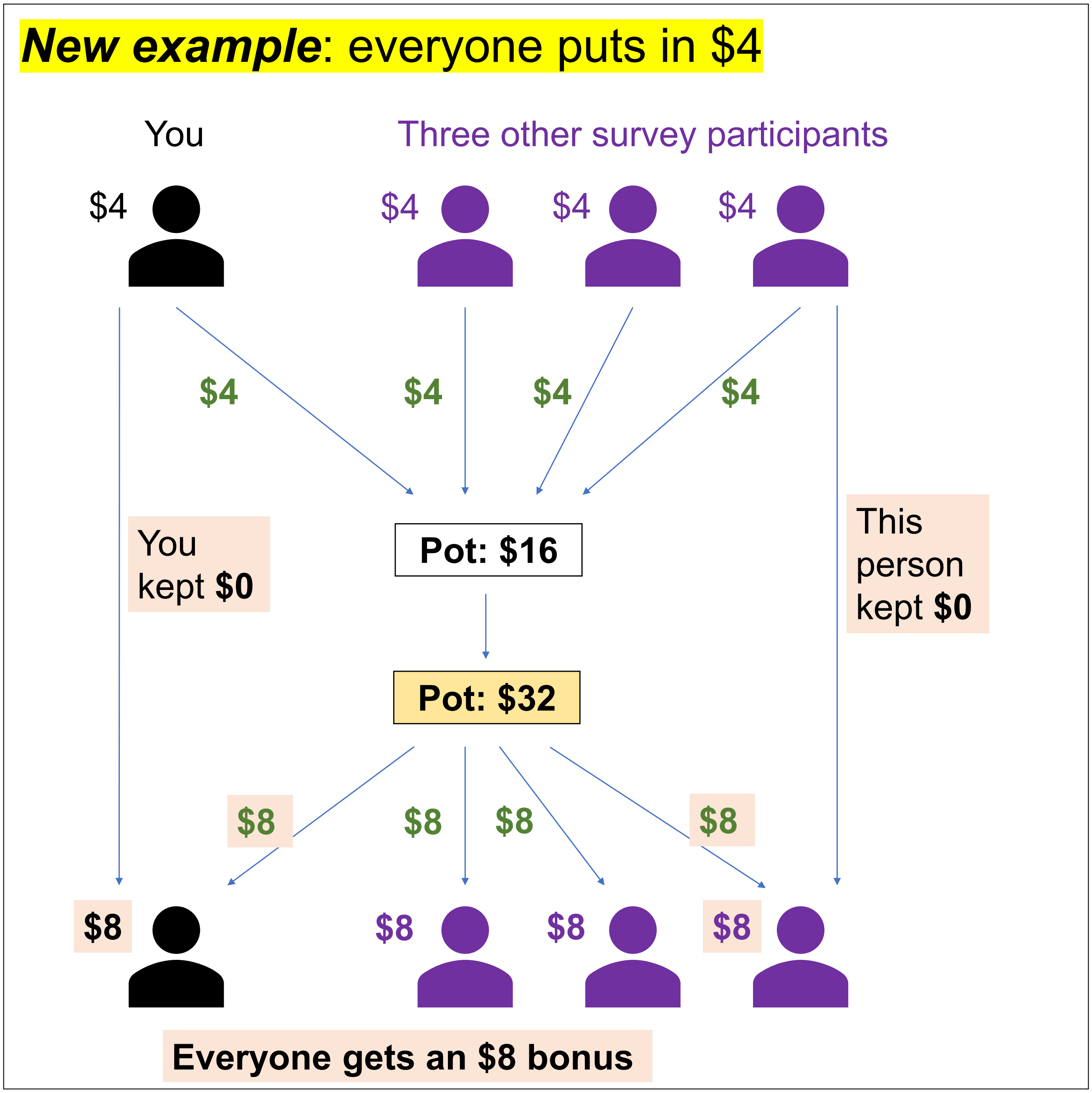
New example: everyone puts in $4
You
Three other survey participants
$4
$4
$4
$4
$4
$4
$4
$4
You kept $0
Pot: $16
This person kept $0
Pot: $32
$8
$8
$8
$8
$8
$8
$8
$8
Everyone gets an $8 bonus

[Page Break]

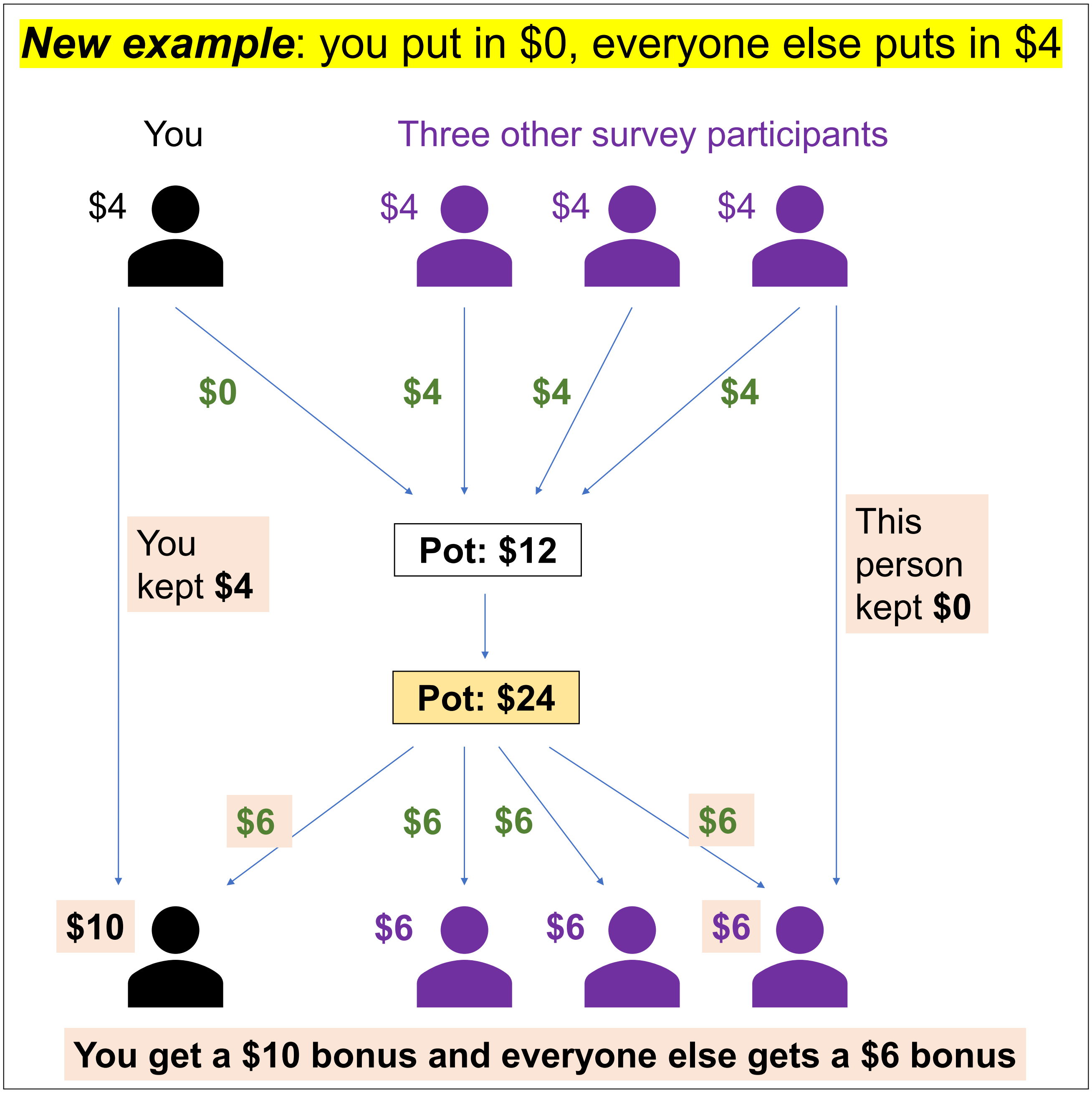


[Page Break]

[ChoiceB] **Scenario B**

In this scenario, you will be matched with three other participants who each made their own decision of how much to put in the pot. Each person's bonus is determined by the decisions of all four participants.

- Put nothing into the pot. Keep all $4 to myself. (0)
- Put $0.50 into the pot. Keep $3.50 to myself. (0.5)
- Put $1.00 into the pot. Keep $3.00 to myself. (1)
- Put $1.50 into the pot. Keep $2.50 to myself. (1.5)

- Put $2.00 into the pot. Keep $2.00 to myself. (2)
- Put $2.50 into the pot. Keep $1.50 to myself. (2.5)
- Put $3.00 into the pot. Keep $1.00 to myself. (3)
- Put $3.50 into the pot. Keep $0.50 to myself. (3.5)
- Put all $4 into the pot. Keep nothing to myself. (4)

[Page Break]

###### 4.2.5 ScenarioCDEF

[ChoiceC] **Scenario C (3 of 6)**

In this scenario, you are matched with one survey participant.

You choose **green** or **orange**.
The other participant also chooses **green** or **orange**.

If they pick green and you pick green: you get $4.00 and they get $4.00.
If they pick green and you pick orange: you get $3.50 and they get $2.00.
If they pick orange and you pick green: you get $2.00 and they get $3.50.
If they pick orange and you pick orange: you get $1.50 and they get $1.50.

It may be useful to consider the other participant's perspective to think about what they might do. This may help:

If you pick green and they pick green: you get $4.00 and they get $4.00.
If you pick green and they pick orange: you get $2.00 and they get $3.50.
If you pick orange and they pick green: you get $3.50 and they get $2.00.
If you pick orange and they pick orange: you get $1.50 and they get $1.50.

- I choose GREEN (1)
- I choose ORANGE (0)

[Page Break]

[ChoiceD] **Scenario D (4 of 6)**

In this scenario, you are matched with one survey participant.

You choose **green** or **orange**.
The other participant also chooses **green** or **orange**.

If they pick green and you pick green: you get $4.00 and they get $4.00.
If they pick green and you pick orange: you get $1.50 and they get $4.50.
If they pick orange and you pick green: you get $4.50 and they get $1.50.
If they pick orange and you pick orange: you get $2.00 and they get $2.00.

It may be useful to consider the other participant's perspective to think about what they might do. This may help:

If you pick green and they pick green: you get $4.00 and they get $4.00.
If you pick green and they pick orange: you get $4.50 and they get $1.50.

If you pick orange and they pick green: you get $1.50 and they get $4.50.
If you pick orange and they pick orange: you get $2.00 and they get $2.00.

- I choose GREEN (1)
- I choose ORANGE (0)

[Page Break]

[ChoiceE] **Scenario E (5 of 6)**

In this scenario, you are matched with one survey participant.

You choose **green** or **orange**.
The other participant also chooses **green** or **orange**.

If they pick green and you pick green: you get $3.00 and they get $3.00.
If they pick green and you pick orange: you get $4.00 and they get $0.25.
If they pick orange and you pick green: you get $0.25 and they get $4.00.
If they pick orange and you pick orange: you get $1.00 and they get $1.00.

It may be useful to consider the other participant's perspective to think about what they might do. This may help:

If you pick green and they pick green: you get $3.00 and they get $3.00.
If you pick green and they pick orange: you get $0.25 and they get $4.00.
If you pick orange and they pick green: you get $4.00 and they get $0.25.
If you pick orange and they pick orange: you get $1.00 and they get $1.00.

- I choose GREEN (1)
- I choose ORANGE (0)

[Page Break]

[ChoiceF] **Scenario F (6 of 6)**

In this scenario, you are matched with one survey participant.

You choose **green** or **orange**.
The other participant also chooses **green** or **orange**.

If they pick green and you pick green: you get $3.00 and they get $3.00.
If they pick green and you pick orange: you get $3.50 and they get $1.00.
If they pick orange and you pick green: you get $1.00 and they get $3.50.
If they pick orange and you pick orange: you get $1.25 and they get $1.25.

It may be useful to consider the other participant's perspective to think about what they might do. This may help:

If you pick green and they pick green: you get $3.00 and they get $3.00.
If you pick green and they pick orange: you get $1.00 and they get $3.50.
If you pick orange and they pick green: you get $3.50 and they get $1.00.
If you pick orange and they pick orange: you get $1.25 and they get $1.25.

- I choose GREEN (1)
- I choose ORANGE (0)

[Page Break]

###### 4.2.6 SocialSurvey

[SocialSurveyIntro] **Social questionnaire**

Now please answer these 10 questions, then you're done!

[Q0] Helping others is usually a waste of time.

- Strongly agree (5)
- Somewhat agree (4)
- Neither agree nor disagree (3)
- Somewhat disagree (2)
- Strongly disagree (1)

[Q1] When given the opportunity, I enjoy aiding others who are in need.

- Strongly agree (5)
- Somewhat agree (4)
- Neither agree nor disagree (3)
- Somewhat disagree (2)
- Strongly disagree (1)

[Q2] Helping friends and family is one of the great joys in life.

- Strongly agree (5)
- Somewhat agree (4)
- Neither agree nor disagree (3)
- Somewhat disagree (2)
- Strongly disagree (1)

[Q3] It feels wonderful to assist others in need.

- Strongly agree (5)
- Somewhat agree (4)
- Neither agree nor disagree (3)
- Somewhat disagree (2)
- Strongly disagree (1)

[Q4] Volunteering to help someone is very rewarding.

- Strongly agree (5)
- Somewhat agree (4)
- Neither agree nor disagree (3)
- Somewhat disagree (2)
- Strongly disagree (1)

[Q5] I dislike giving directions to strangers who are lost.

- Strongly agree (5)

- Somewhat agree (4)
- Neither agree nor disagree (3)
- Somewhat disagree (2)
- Strongly disagree (1)

[Q6] Unless they are part of my family, helping the elderly isn't my responsibility.

- Strongly agree (5)
- Somewhat agree (4)
- Neither agree nor disagree (3)
- Somewhat disagree (2)
- Strongly disagree (1)

[Q7] If the person in front of me in the check-out line at a store was a few cents short, I would pay the difference.

- Strongly agree (5)
- Somewhat agree (4)
- Neither agree nor disagree (3)
- Somewhat disagree (2)
- Strongly disagree (1)

[Q8] I plan to donate my organs when I die with the hope that they will help someone else live.

- Strongly agree (5)
- Somewhat agree (4)
- Neither agree nor disagree (3)
- Somewhat disagree (2)
- Strongly disagree (1)

[Q9] Helping people does more harm than good because they come to rely on others and not themselves.

- Strongly agree (5)
- Somewhat agree (4)
- Neither agree nor disagree (3)
- Somewhat disagree (2)
- Strongly disagree (1)
